# ApoER2-Dab1 disruption as the origin of pTau-related neurodegeneration in sporadic Alzheimer’s disease

**DOI:** 10.1101/2023.05.19.23290250

**Authors:** Christopher E. Ramsden, Daisy Zamora, Mark S. Horowitz, Jahandar Jahanipour, Gregory S. Keyes, Xiufeng Li, Helen C. Murray, Maurice A. Curtis, Richard M. Faull, Andrea Sedlock, Dragan Maric

**Affiliations:** Lipid Peroxidation Unit, Laboratory of Clinical Investigation, National Institute on Aging, NIH 251 Bayview Blvd., Baltimore, MD, 21224, USA; Intramural Program of the National Institute on Alcohol Abuse and Alcoholism, NIH, Bethesda, MD, 20892, USA; Department of Physical Medicine and Rehabilitation, School of Medicine, University of North Carolina, Chapel Hill, NC, 27599, USA; Flow and Imaging Cytometry Core Facility, National Institute of Neurological Disorders and Stroke, NIH, Bethesda, MD, 20892, USA; Department of Anatomy and Medical Imaging and Centre for Brain Research, Faculty of Medical and Health Science, University of Auckland, Private Bag, Auckland, 92019, New Zealand; Laboratory of Functional and Molecular Imaging, National Institute of Neurological Disorders and Stroke, NIH, Bethesda, MD, 20892, USA

**Author notes:** Corresponding author: Christopher Ramsden, MD, PhD., Chief, Lipid Peroxidation Unit, Laboratory of Clinical Investigation, NIA/NIH, Baltimore, MD, 21224.

## Abstract

**BACKGROUND:** Sporadic Alzheimer’s disease (sAD) is not a global brain disease. Specific regions, layers and neurons degenerate early while others remain untouched even in advanced disease. The prevailing model used to explain this selective neurodegeneration—prion-like Tau spread—has key limitations and is not easily integrated with other defining sAD features. Instead, we propose that in humans Tau hyperphosphorylation occurs locally via disruption in ApoER2-Dab1 signaling and thus the presence of ApoER2 in neuronal membranes confers vulnerability to degeneration. Further, we propose that disruption of the Reelin/ApoE/ApoJ-ApoER2-Dab1 P85α-LIMK1-Tau-PSD95 (RAAAD-P-LTP) pathway induces deficits in memory and cognition by impeding neuronal lipoprotein internalization and destabilizing actin, microtubules, and synapses. This new model is based in part on our recent finding that ApoER2-Dab1 disruption is evident in entorhinal-hippocampal terminal zones in sAD. Here, we hypothesized that neurons that degenerate in the earliest stages of sAD (1) strongly express ApoER2 and (2) show evidence of ApoER2-Dab1 disruption through co-accumulation of multiple RAAAD-P-LTP components.

**METHODS:** We applied *in situ* hybridization and immunohistochemistry to characterize ApoER2 expression and accumulation of RAAAD-P-LTP components in five regions that are prone to early pTau pathology in 64 rapidly autopsied cases spanning the clinicopathological spectrum of sAD.

**RESULTS:** We found that: (1) selectively vulnerable neuron populations strongly express ApoER2; (2) numerous RAAAD P-LTP pathway components accumulate in neuritic plaques and abnormal neurons; and (3) RAAAD-P-LTP components were higher in MCI and sAD cases and correlated with histological progression and cognitive deficits. Multiplex-IHC revealed that Dab1, pP85α_Tyr607_, pLIMK1_Thr508_, pTau and pPSD95_Thr19_ accumulated together within dystrophic dendrites and soma of ApoER2-expressing neurons in the vicinity of ApoE/ApoJ enriched extracellular plaques. These observations provide evidence for molecular derangements that can be traced back to ApoER2-Dab1 disruption, in each of the sampled regions, layers, and neuron populations that are prone to early pTau pathology.

**CONCLUSION:** Findings support the RAAAD-P-LTP hypothesis, a unifying model that implicates dendritic ApoER2-Dab1 disruption as the major driver of both pTau accumulation and neurodegeneration in sAD. This model provides a new conceptual framework to explain why specific neurons degenerate and identifies RAAAD-P-LTP pathway components as potential mechanism-based biomarkers and therapeutic targets for sAD.

## Introduction

Sporadic Alzheimer’s disease (sAD) is not a global brain disease. Specific regions, layers and neurons degenerate early while others remain untouched even in advanced disease.^1–4^ The quest to unravel the molecular basis of this selective degeneration started more than a century ago and has centered on hyper-phosphorylated forms of the microtubule associated protein Tau (pTau) for decades. pTau accumulates within four sAD lesions—neuropil threads (NTs), neurofibrillary tangles (NFTs), neuritic plaques (NPs), and granulovacuolar degeneration bodies (GVDs). The molecular events underlying pTau accumulation and the mechanistic links between NTs, NFTs, NPs, and GVDs are incompletely understood. Among these pTau-containing pathologies, NFTs are most often used to track and distinguish different pathological stages of sAD due to their remarkably consistent anatomical site(s) of origin and sequence of progression throughout the brain (**Fig 1**), and strong correlation with memory loss.^4–6^ In this manuscript we focus on five neuron populations that accumulate pTau in the earliest stages of sAD (**Fig 1**).^1–3 7–13^ The identification of a single shared mechanism that renders these neurons selectively vulnerable to pTau-related neurodegeneration—and provides a missing molecular link underlying the genesis of NTs, NFTs, NPs, and GVDs—could have major implications for the prevention and treatment of sAD.

**Fig 1:**
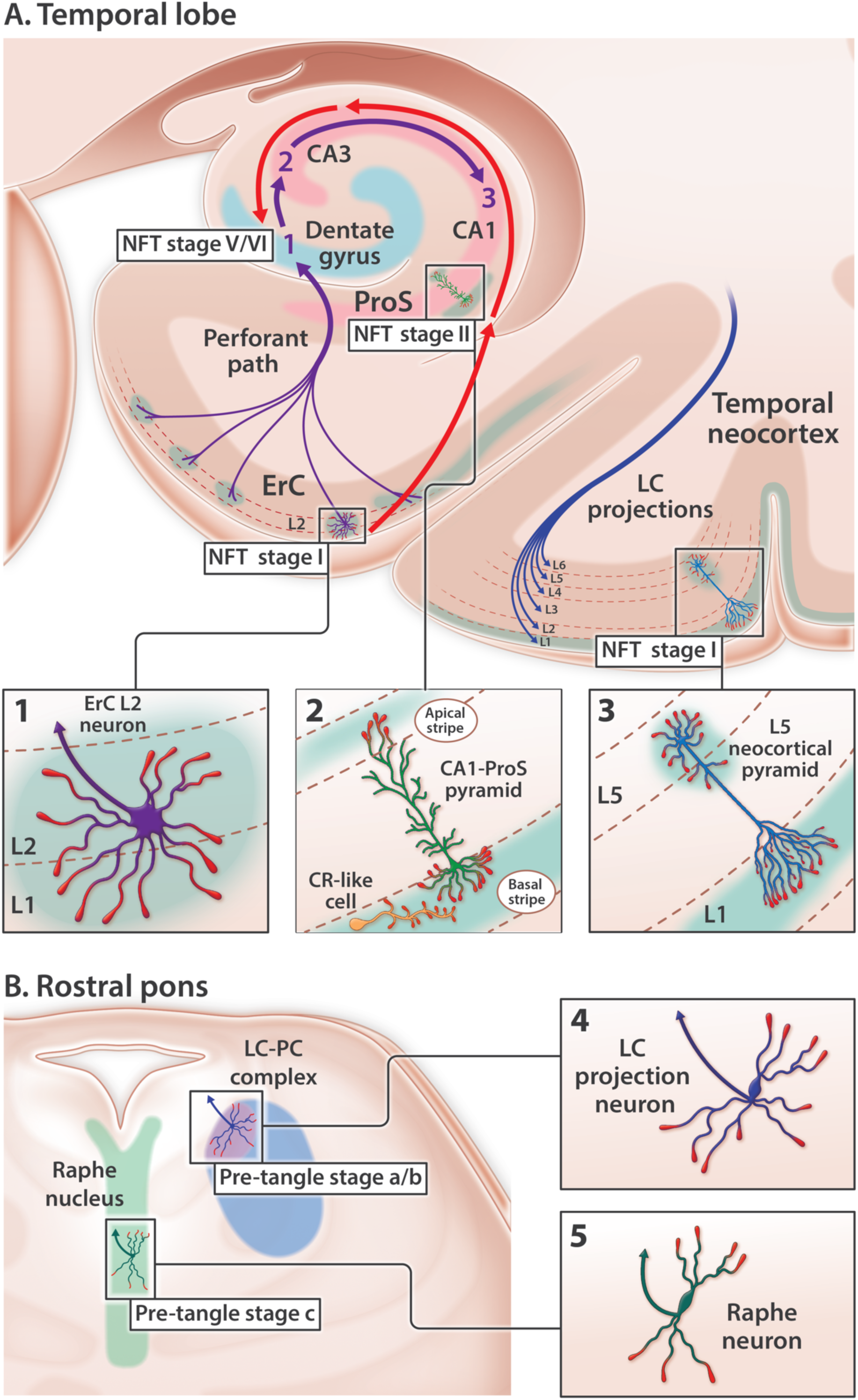
Anatomical sites of origin and ordered progression of NFT pathology in early sAD. pTau accumulates in five neuron populations within the temporal lobe (**A**) and rostral pons (**B**) in the earliest stages of sAD. (**A**) Temporal lobe: red arrows trace the stereotypical progression of pTau pathology in the medial temporal lobe memory system. In NFT stage I, pTau pathology is confined to ErC L2 projection neurons. In NFT stage II, pTau pathology progresses to include the basal lamina of the ProS-CA1 border region. In successive NFT stages, pTau pathology progresses throughout the cornu ammonis. Dentate granule neurons (light blue arc) are spared from NFT pathology until late-stage sAD (NFT stages V-VI). Purple arrows denote the unidirectional tri-synaptic circuitry underlying memory formation. The contrasting directionality of red and purple arrows indicates that NFT pathology progresses in a direction that is opposite to the synaptic connections underlying memory. pTau pathology is also evident in solitary L5 and L3 pyramids in the temporal neocortex very early in sAD (NFT stage I). (**B**) Rostral pons: pTau pathology classically begins in the LC-PC complex (pre-tangle stage a/b) before progressing to include the raphe nucleus (pre-tangle stage c) and ErC L2 (NFT stage I, see **A**). LC axonal projections (blue arrow in **B4**) innervate virtually the entire brain including all layers of the temporal neocortex (depicted by blue axon terminals in L1-L6 of the temporal cortex). Raphe neurons (green arrow in **B5**) also project widely throughout the brain. LC and Raphe neurons therefore do not selectively innervate ErC L2.

### Rationale and limitations of the Tau prion-like connectome-based spread model for NFT progression

The prevailing model cited to account for the ordered sequence of NFT progression posits that prion-like properties enable pathogenic forms of Tau to spread from donor neurons to recipient neurons and to convert normal Tau to pathogenic Tau (**Fig 2A** and **Suppl Table 1**).^11 14–17^ This ‘prion-like connectome-based spread’ hypothesis provides the rationale for a new class of immunotherapeutics targeting extracellular Tau.^18 19^ While rodent and cellular models have shown that trans-synaptic Tau transmission is possible,^20^ three pivotal observations in humans contradict a simple model of connectome-based spread (reviewed in **Suppl Table 1**). First, NFT progression proceeds in a direction opposite to unidirectional connectivity in the medial temporal lobe memory system (**Fig 1A**).^21 22^ Second, pTau pathology classically originates in the locus coeruleus (LC) before progressing specifically to L2 of the entorhinal region (ErC L2).^9 14^ However, LC neurons project widely throughout the brain and do not selectively innervate ErC L2 (**Fig 1B**).^23–27^ Third, individual projection neurons classically innervate hundreds or even thousands of neighboring target neurons ^28–31^. Yet in the earliest stages of sAD pTau accumulates in rare, solitary L5 and L3 neocortical pyramids while sparing neighboring neurons (**Fig 1A**). ^11 32^ Accommodation of the prion-like connectome-based spread hypothesis therefore requires speculation about connections that are not yet known to exist ^7^ and a revised model of brain connectivity (reviewed in **Suppl Table 1**).^9 10^ Further, while the Tau prion-like hypothesis makes predictions about spread it does not attempt to explain how the first lesions develop.

**Fig 2:**
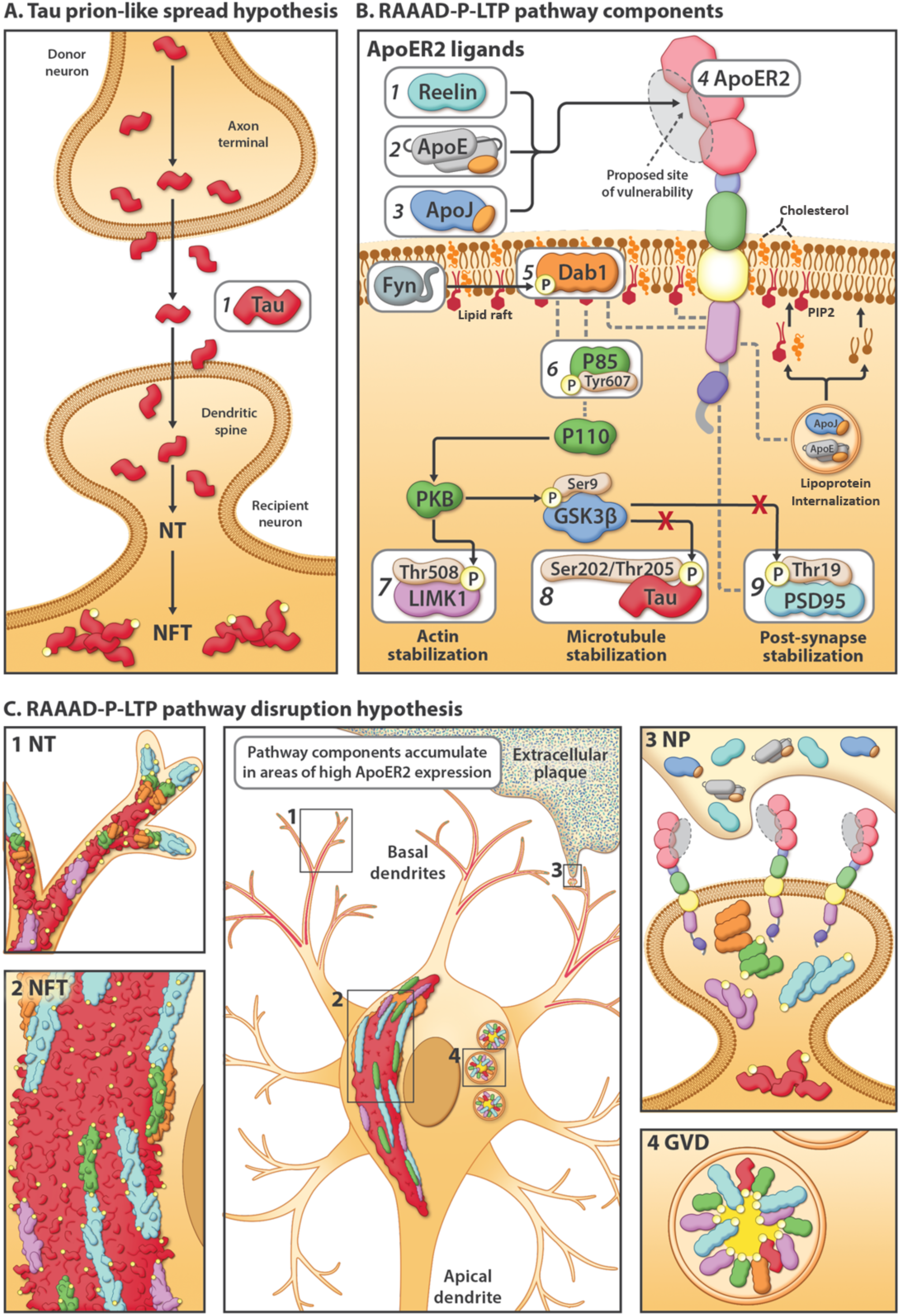
Competing explanatory hypotheses for pTau pathology in sAD: Prion-like connectome-based spread versus RAAAD-P-LTP pathway disruption. (**A**) The prion-like spread hypothesis focuses exclusively on Tau (**A1**) and predicts a connectome-based sequence of NFT progression. (**B**) The RAAAD-P-LTP pathway includes 9 core components (**B1-9**) that mediate cytoskeletal and synaptic changes underlying memory. Under physiological conditions, binding of ligands (Reelin [**B1**]), ApoE [**B2**]), ApoJ [**B3**]) to ApoER2 [**B4**] triggers recruitment of Dab1 [**B5**] and P85α [**B6**] to lipid rafts, formation of ApoER2-Dab1-P85α signaling complexes, and phosphorylation, activation, and degradation of Dab1. Ensuing activation of the Dab1-P85α/PI3K cascade evokes phosphorylation of LIMK1 [**B7**] to stabilize the actin cytoskeleton, and inhibition of GSK3β, which in turn inhibits phosphorylation of Tau [**B8**] and PSD95 [**B9**] to stabilize microtubules and postsynaptic receptor complexes, respectively. ApoE receptor-mediated lipoprotein internalization supplies the lipid cargo required to shape and remodel synaptic membranes, including the lipid rafts that harbor ApoER2-Dab1-P85α complexes. (**C**) The RAAAD-P-LTP pathway disruption hypothesis can mechanistically & spatially link four pTau containing lesions (NTs [**C1**], NFTs [**C2**], NPs [**C3**], GVDs [**C4**]). ApoER2-Dab1 disruption is proposed to trigger Tau hyperphosphorylation and thus the presence of ApoER2 in neuronal membranes confers vulnerability to pTau-related degeneration. In this model, a triggering molecular lesion or competitive inhibition among ligands disrupts ApoER2 binding leading to extracellular accumulation of ApoER2 ligands in NPs. Impaired ApoER2 signaling disrupts Dab1 degradation leading to localized Dab1 accumulation. Ensuing disruption of the Dab1-P85α/PI3K-LIMK1 arm destabilizes the actin cytoskeleton and induces pP85α_Tyr607_ and pLIMK1_Thr508_ accumulation. Impaired Reelin-ApoER2-Dab1-P85α/PI3K signaling activates GSK3β; ensuing phosphorylation of Tau and PSD95 destabilizes microtubules and postsynaptic receptor complexes, and promotes accumulation of pTau and pPSD95 in NTs, NFTs, NPs and GVDs. Thus, deficits in memory and cognition in sAD are attributed to impaired neuronal lipoprotein internalization and destabilization of actin, microtubules, and synapses.

### Dendritic RAAAD-P-LTP pathway disruption as an alternative explanation for pTau pathology

In rodent and cellular models, ApoER2 has been shown to play central roles in cytoskeletal and synaptic plasticity ^33–36^ and learning and memory,^35 37–39^ and to regulate Tau phosphorylation ^40–44^ as part of a signaling cascade involving its extracellular ligands (Reelin,^44^ ApoE,^43^ ApoJ ^45^) and intraneuronal signaling partners (Dab1, P85α/PI3K, LIMK1, Tau, PSD95) (**Fig 2B**)(reviewed in ^46^). Despite these crucial functions in rodents, comparatively little is known about the functions and distribution of ApoER2 in the human brain. Here, we propose and provide evidence supporting dendritic <u>R</u>eelin/<u>A</u>poE/<u>A</u>poJ-<u>A</u>poER2-<u>D</u>ab1-<u>P</u>85α/PI3K-<u>L</u>IMK1-<u>T</u>au <u>P</u>SD95 (RAAAD-P-LTP) pathway disruption as a new, alternative model that could explain both the anatomical site(s) of origin and progression of pTau-associated neurodegeneration in human sAD, and provide missing molecular links between NTs, NFTs, NPs, and GVDs (**Fig 2B-C** and **Suppl Table 1**). The RAAAD-P-LTP disruption hypothesis has three central tenets: (1) disruption of ApoER2-Dab1 in dendritic spines induces local Tau hyperphosphorylation and pTau accumulation; (2) pTau accumulation is only one of numerous neurodegenerative pathologies triggered by ApoER2-Dab1 disruption; and (3) neurons with high ApoER2 expression and demand for RAAAD-P-LTP pathway activation are particularly vulnerable to pTau-related degeneration. This hypothesis is based in part upon our recent findings of pathological accumulation of ApoER2 together with its extracellular ligands (ApoE and Reelin), intracellular adaptor protein (Dab1), and downstream ApoER2-Dab1 signaling partners (pP85α_Tyr607_, pLIMK1_Thr508_, pTau, and pPSD95_Thr19_) within entorhinal-hippocampal ‘perforant path’ terminal zones in human sAD.^46^ However, these perforant path terminal zones develop pTau pathology in the later stages of sAD (**Fig 1**) and thus are not optimal for addressing the origin. The purpose of the present study is therefore to characterize ApoER2 expression and to search for evidence of RAAAD-P-LTP pathway disruption in and around five neuron populations that accumulate pTau in the earliest stages of sAD (**Fig 1**): ErC L2 neurons, basal ProS-CA1 pyramids, solitary L5 and L3 neocortical pyramids, and pontine LC and raphe nucleus neurons. Based on the RAAAD-P-LTP model we hypothesized that (1) these five vulnerable neuron populations strongly express ApoER2; and (2) multiple RAAAD-P-LTP pathway components accumulate inside and in the immediate vicinity of these vulnerable neurons in Mild Cognitive Impairment (MCI) and sAD cases.

Using single-marker immunohistochemistry (IHC), *in situ* hybridization (ISH), and multiplex fluorescence-IHC (MP-IHC) to examine postmortem specimens from 64 cases spanning the clinicopathological spectrum of sAD (**Table 1**) we observed: (1) striking laminar and cellular distributions of ApoER2 with strong expression in neuron populations known to accumulate pTau early in sAD and lower or absent expression in neuron populations that are spared from NFT pathology; and (2) accumulations of extracellular plaque associated ApoER2 ligands (ApoE, ApoJ) and intraneuronal ApoER2 signaling partners (Dab1, pP85α_Tyr607_, pLIMK1_Thr508_, pPSD95_Thr19_, pTau_Ser202/Thr205_), within these same regions and layers in MCI and sAD cases that were absent or less pronounced in non-AD controls. MP-IHC revealed that pTau accumulates together with Dab1, pP85α_Tyr607_, pLIMK1_Thr508_, and pPSD95_Thr19_ within MAP2-labeled dystrophic dendrites and soma of NFT and/or GVD-bearing ApoER2-expressing neurons and in the vicinity of ApoE/ApoJ-enriched NPs. These observations provide evidence for both upstream and downstream molecular derangements that can be traced back to dendritic ApoER2-Dab1 disruption, in each of the sampled regions, layers, and neuron populations that are prone to early development of NFT pathology. Collective findings reveal that pTau is only one of many RAAAD-P-LTP pathway components that accumulate in multiple neuroanatomical sites in the early stages of sAD and provide support for the concept that dendritic ApoER2-Dab1 disruption is a major driver of degeneration and pTau accumulation in selectively vulnerable neurons in humans.

**Table 1.** Summary characteristics (n=64)

|  | Banner (n=34) <sup>a</sup> |  |  |  | New Zealand (n=18) <sup>b</sup> |  | UK-ADRC (n=12) |  |  |
| --- | --- | --- | --- | --- | --- | --- | --- | --- | --- |
|  | AD<br>(n=10) | MCI<br>(n=9) | Control<br>(n=10) | Young<br>Control<br>(n=5) | AD<br>(n=10) | Control<br>(n=8) | AD<br>(n=4) | MCI<br>(n=4) | Control<br>(n=4) |
| **Demographic and clinical characteristics** |  |  |  |  |  |  |  |  |  |
| Age, y, median (range) | 82.5<br>(73-90) | 85<br>(76-90) | 84<br>(71-90) | 52<br>(38-61) | 79<br>(60-94) | 73<br>(59-78) | 84.5<br>(75-91) | 85.5<br>(84-92) | 84.5<br>(71-92) |
| Female, n | 6 | 6 | 4 | 2 | 4 | 1 | 2 | 2 | 2 |
| PMI | 3.5<br>(2.2-4.9) | 3.2<br>(2-4.2) | 3<br>(2.1-4.6) | 3.1<br>(2.3-4.7) | 12<br>(5-24) | 17<br>(5.5-25) | 3<br>(2.2-3.3) | 2.6<br>(2.3-3.5) | 2.7<br>(1.3-4) |
| MMSE (0-30), median<br>(range) | 13.5<br>(2-24) | 27<br>(22-29) | 28<br>(27-30) |  |  |  | 3.5<br>(0-10) | 26.5<br>(24-28) | 28<br>(24-30) |
| **Neuropathology and genetics** |  |  |  |  |  |  |  |  |  |
| NFT stage (0-6), median<br>(range) | 6<br>(5-6) | 4<br>(1-4) | 1<br>(1-3) | 1<br>(0-1) | 5<br>(3-6) | 1.5<br>(1-3) | 6<br>(6-6) | 3.5<br>(2-4) | 1<br>(0-2) |
| Neuritic plaque density (0-3), median (range) | 3<br>(3-3) | 3<br>(0-3) | .5<br>(0-1) | 0<br>(0-1) | 2<br>(1-3) |  | 3<br>(3-3) | 0<br>(0-2) | 0<br>(0-0) |
| Amyloid plaques (0-15),<br>median (range) | 15<br>(12.5-15) | 8.25<br>(0-13.5) | .25<br>(0-6.5) | 0<br>(0-1) |  |  |  |  |  |
| ApoE, n |  |  |  |  |  |  |  |  |  |
| 3/3 | 5 | 4 | 7 | 5 | 4 | 7 | 1 | 2 | 3 |
| 2/3 | 1 | 5 | 1 | 0 | 0 | 0 | 1 | 0 | 0 |
| 3/4 | 4 | 0 | 2 | 0 | 5 | 0 | 1 | 2 | 1 |
| 4/4 | 0 | 0 | 0 | 0 | 1 | 0 | 1 | 0 | 0 |

<sup>a</sup> Cases that were ≥90 years of age were classified as age 90. Scores for amyloid plaques include the entorhinal, hippocampus, temporal, parietal, and frontal areas. Each was scored according to the CERAD templates <sup>59</sup> using Campbell-Switzer silver stain, Gallyas silver stain and Thioflavin S stains. Not all tissues were available for all cases.
<sup>b</sup> MMSE scores for both groups were not available. Control group was not evaluated for neuritic plaque density or amyloid plaques.

## MATERIALS AND METHODS

### Study cohorts/Postmortem specimens

#### Brain and Body Donation Program (BBDP) at the Banner Sun Health Research Institute

Rapidly-autopsied, minimally-fixed specimens from 34 BBDP cases spanning the clinicopathological spectrum of sAD (**Table 1 & Suppl Table 2**) acquired from the BBDP at the Banner Sun Health Research Institute (http://www.brainandbodydonationprogram.org) ^46^ were used for the main analyses of ErC, ProS-CA1 border region and temporal neocortex. Briefly, 6μm-thick formalin-fixed, paraffin-embedded (FFPE) tissue sections containing (1) ventral ErC at the level of the amygdala; (2) medial temporal lobe at the level of the body of the hippocampus; and (3) middle temporal gyrus, were obtained from BBDP. BBDP employed a rapid on-call autopsy team to achieve short PMI to mitigate limitations due to tissue degradation. Standardized fixation procedures were employed using 1 cm^3^ tissue blocks fixed in 10% neutral buffered formalin (NBF) for 48 hrs. All BBDP subjects provided written consent for study procedures, autopsy and sharing of de-identified data prior to enrollment. The study and its consenting procedures were approved by the Western Institutional Review Board (IRB) of Puyallup, Washington. The Banner BBDP population has been extensively described.^47–50^ Most BBDP donors were enrolled as cognitively normal volunteers residing in the retirement communities near Phoenix, AZ, USA. Following provision of informed consent, donors received standardized medical, neurological, and neuropsychological assessments during life. Neuropathological and cognitive endpoints captured by BBDP are detailed in a previous publication.^47^ NFT stage (0-6), Total amyloid plaques (0-15), Neuritic plaque density (0-3), and *APOE* status were used in the present study (**Table 1 & Suppl Table 2**). The antemortem Mini-Mental Status Exam (MMSE, 0-30) test had the least missing data and was the main cognitive endpoint of for the present study.

#### University of Auckland, New Zealand Neurological Foundation Human Brain Bank (UA-HBB) ^51^

Minimally-fixed specimens from 10 sAD cases and 8 neurologically normal control cases (**Table 1 & Suppl Table 2**) acquired from the UA-HBB and the Human Anatomy Laboratory within the Department of Anatomy and Medical Imaging at the University of Auckland^51 52^ were used for the main analyses of upper pons (LC plus raphe nucleus) and as a validation cohort for pathologies observed in the ErC and ProS-CA1 regions in BBDP specimens. Briefly, 5μm-thick FFPE tissue sections including (1) transverse sections of the upper pons at the level of the LC and (2) medial temporal lobe specimens at the level of the body of the hippocampus were obtained. The UA-HBB team carefully dissected the upper pons and each block was visually inspected the presence of the LC and raphe nucleus. ^52^ Tissue was donated with informed consent from the family before brain removal; all procedures were approved by the University of Auckland Human Participants Ethics Committee (Ref: 011654). The right hemisphere of each brain was either perfusion or immersion fixed (depending on the location of the mortuary conducting the brain removal) in 15% formaldehyde in 0.1 M phosphate buffer for 48 h at room temperature. The upper pons block was paraffin-embedded in a transverse orientation as previously described. The sAD cases had a clinical history of dementia, and the clinical AD diagnosis was confirmed by independent pathological assessment. The neurologically normal control cases had no history of neurological abnormalities, and no substantial neuropathology was noted upon autopsy. Neuropathological endpoints have been extensively described.^51 52^ NFT stage (0-6), Thal phase (0-5), Neuritic plaque density (0-3), and *APOE* status were used in the present study (**Table 1**).

#### University of Kentucky Alzheimer’s Disease Research Center (UK-ADRC) Biobank

FFPE blocks containing the superior and middle temporal gyrus (SMTG) from 12 rapidly-autopsied cases spanning the clinicopathological spectrum of sAD acquired through the UK-ADRC Biobank (**Table 1 & Suppl Table 2**) were used as a validation cohort for pathologies observed in BBDP and UA-HBB specimens. Standardized fixation procedures were employed with brain fixed in 10% NBF for approximately one month. UK-ADRC subjects provided written consent for study procedures, autopsy and sharing of de-identified data prior to enrollment. The study and its consenting procedures were approved by the IRB (44009) of the University of Kentucky. Neuropathological and cognitive endpoints captured by UK-ADRC have been extensively described.^53^ NFT stage (0-6), Thal phase (0-5), Neuritic plaque density (0-3), MMSE (0-30), and *APOE* status were used in the present study (**Table 1**).

#### Single-marker immunohistochemistry (IHC)

Single-marker IHC was performed by Histoserv (Gaithersburg, MD, USA) and in our NIA/NIAAA laboratory as previously described.^46^ Briefly, combined blocks containing temporal cortex from sAD cases and non-AD controls were sectioned into 6μm-thick coronal sections. Optimal immunostaining conditions were then empirically determined using an incremental heat induced epitope retrieval (HIER) method using 10 mM Na/Citrate pH6 and/or 10 mM TRIS/EDTA pH9 buffer that were heated at 70°C for 10-40min. Selected antibodies with suboptimal HIER treatment underwent additional retrieval rounds using formic acid (88%, 10-20 min) or proteinase K (Dako S3020) for 5 min at room temperature (RT). Secondary antibodies (Jackson ImmunoResearch) used for single IHC chromogenic staining were appropriately matched to the host class/subclass of the primary antibody. These combined sAD plus anatomically-matched control tissue blocks were used to generate negative controls (comparing staining results versus pre-immune serum and with the primary antibody omitted) and positive controls (comparing staining results in temporal or frontal cortex specimens from confirmed AD cases with known region-specific Aβ and tangle scores versus non-AD controls). These empirically determined optimal conditions were then used to immunolabel sets of slides using IHC and MP-IHC (as described below). Briefly, for IHC, sections were first deparaffinized using standard Xylene/Ethanol/Rehydration protocol followed by antigen unmasking with 10–40 min HIER at 70°C or formic acid for 10-20 min at RT, as described above. The antibodies selected for this study have previously been used for IHC in human tissues and western blot assays (see **Suppl Table 3** for antibody sources and technical specifications). The same IHC-validated antibodies were used to immunolabel cytoarchitectural markers, core components of the RAAAD-P-LTP pathway, and hallmark AD pathologies in a recently published a manuscript ^46^ in human brain.

#### Multiplex fluorescence immunohistochemistry and in situ hybridization

Multiplex fluorescence immunohistochemistry (MP-IHC) and multiplex fluorescence *in situ* hybridization (MP-ISH) were performed on 5μm-thick or 6μm-thick sections that were mounted on Leica Apex Superior adhesive slides (VWR, 10015-146) or UberFrost (InstrumeC) slides to prevent tissue loss. We completed one round of MP-ISH and up to 6 iterative rounds of sequential MP-IHC staining with select mRNA probes and antibody panels targeting the RAAAD-P-LTP pathway and/or classical sAD biomarkers, alongside classical cytoarchitectural biomarkers to map brain tissue parenchyma (see **Suppl Table 3**). MP-ISH was carried out on a subset of sections using a custom designed RNA probes and standard RNAScope Multiplex Fluorescent V2 hybridization kits (Advanced Cell Diagnostics, Inc.), per manufacturer’s instructions. Fluorophore-labelled mRNA/protein targets from each round of staining were imaged by multispectral epifluorescence microscopy followed by mRNA probe/antibody stripping and tissue antibody re-staining steps to repeat the cycle, as previously described,^46 54^ each time using a different antibody panel. Briefly, for screening tissues by MP-ISH/MP-IHC, the sections were first deparaffinized using standard Xylene/Ethanol/Rehydration protocol followed by proteinase and/or HIER target unmasking steps in 10 mM Tris/EDTA buffer for 10 min using an 800W microwave set at 100% power. Select sections were then processed for MP-ISH, as referenced above, and all sections to be sequentially processed for iterative MP-IHC screening were first incubated with Human BD Fc Blocking solution (BD Biosciences) to saturate endogenous Fc receptors, and then in True Black Reagent (Biotium) to quench intrinsic tissue autofluorescence. Sections were then immunoreacted for 1 h at RT using cocktail mixture of immunocompatible antibody panels, with each antibody used at optimal staining concentration, as listed in **Suppl Table 3**. This step was followed by washing off unbound primary antibodies in PBS supplemented with 1 mg/ml bovine serum albumin (BSA) and staining the sections using a 1μg/ml cocktail mixture of the appropriately cross-adsorbed and host and immunoglobulin class/subclass-specific secondary antibodies (purchased from either Thermo Fisher, Jackson ImmunoResearch or Li-Cor Biosciences) conjugated to one of the following spectrally compatible fluorophores: Alexa Fluor 430, Alexa Fluor 488, Alexa Fluor 546, Alexa Fluor 594, Alexa Fluor 647, IRDye 600LT, or IRDye 800CW. After washing off excess secondary antibodies, sections were counterstained using 1μg/ml DAPI (Thermo Fisher Scientific) for visualization of cell nuclei. Slides were then coverslipped using Immu-Mount medium (Thermo Fisher Scientific) and imaged using a multispectral epifluorescence microscope (see below). After imaging, tissue-bound primary and secondary antibodies were both stripped off the slides after a 5 min incubation at RT in NewBlot Nitro 5X Stripping buffer (Li-Cor Biosciences) followed by 1 min additional HIER step in Tris/EDTA buffer. The above processing cycle beginning with re-blocking of tissues in Human BD Fc Blocking solution was repeated and the same sections then incubated using an additional panel of antibodies of interest.

#### Multiplex fluorescence immunohistochemistry image acquisition and computational reconstruction

Images were acquired from MP-ISH and/or MP-IHC probed specimen sections using the Axio Imager.Z2 slide scanning fluorescence microscope (Zeiss) equipped with a 20X/0.8 Plan-Apochromat (Phase-2) non-immersion objective (Zeiss), a high resolution ORCA-Flash4.0 sCMOS digital camera (Hamamatsu), a 200W X-Cite 200DC broad band lamp source (Excelitas Technologies), and 8 customized filter sets (Semrock) optimized to detect the following fluorophores: DAPI, Alexa Fluor 430, Alexa Fluor 488 (or Opal 520), Alexa Fluor 546 (or Opal 570), Alexa Fluor 594 (or Opal 620), Alexa Fluor 647, IRDye 680LT (or Opal 690), and IRDye 800CW. Image tiles (600×600μm viewing area) were individually captured at 0.325 micron/pixel spatial resolution, and the tiles seamlessly stitched into whole specimen images using the ZEN 2 image acquisition and analysis software program (Zeiss), with an appropriate color table having been applied to each image channel to either match its emission spectrum or to set a distinguishing color balance. The RGB histogram of each image was adjusted using Zen software, resulting in optimized signal brightness and contrast, improved dynamic range, exposure correction, and gamma/luminosity-enhancement to reveal hidden/dim image details. The stitched images were exported as tif files, then computationally registered at the subpixel level using affine transformation and corrected for photobleaching, autofluorescence, non-uniform illumination shading, spectral bleed-through, and molecular colocalization artifacts, exported to Adobe Photoshop and overlaid as individual layers to create multicolored contrast-enhanced composite images, as previously described by Maric et al^46 51 55^. Contrast-enhanced composite MP-IHC images were used to provide cytoarchitectural and pathological context for the single-marker IHC images which were used for quantitation.

#### Regional Annotation and Quantitation

Single-marker IHC images were uploaded into HALO 3.3 image analysis software (Indica Labs, Corrales, NM). Boundaries of each anatomical region of interest (ErC, CA1-ProS border region, gray matter of middle temporal gyrus, pontine LC plus raphe nucleus) were identified and annotated using a combination of anatomical landmarks (rhinal sulcus, dentate gyrus, temporal horn of lateral ventricle, pyramidal blades of the dentate gyrus, 4^th^ ventricle) and cytoarchitectonic mapping as previously described.^46^ MP-IHC with immunolabeling of numerous cytoarchitectural markers was performed on serial sections to assist with identification of anatomical landmarks and boundaries including white matter tracts and gray matter-white matter interfaces as previously described. ^46^ Stain positive area as a percentage of each annotated region was quantified using the HALO Area Quantification v2.2.1 module. Plaque-associated objects per mm^2^ within each annotated region were identified and quantified using the HALO Object Colocalization v1.3 module with classifier function enabled as previously described.^46^

#### Statistical analysis

For each annotated region, between-group differences according to each marker were quantified using Kruskal Wallis tests. Variable transformations (e.g., natural log) were used as necessary. A Spearman’s correlation coefficient between each immunohistochemical marker and each AD endpoint (NFT stage, Aβ plaque load, MMSE) was calculated. Graphs showing individual data points in each group and their relationships to sAD endpoints are provided in **Figs 3-4**, **6**, **9**, **10**. In a sensitivity analyses accounting for the false discovery rate, we adjusted the p-values using the two-stage linear step-up procedure described by Benjamini et al.^56 57^ (presented in **Suppl Table 4**). Statistical analyses were conducted in Stata Release 17.^58^

**Fig 3.**
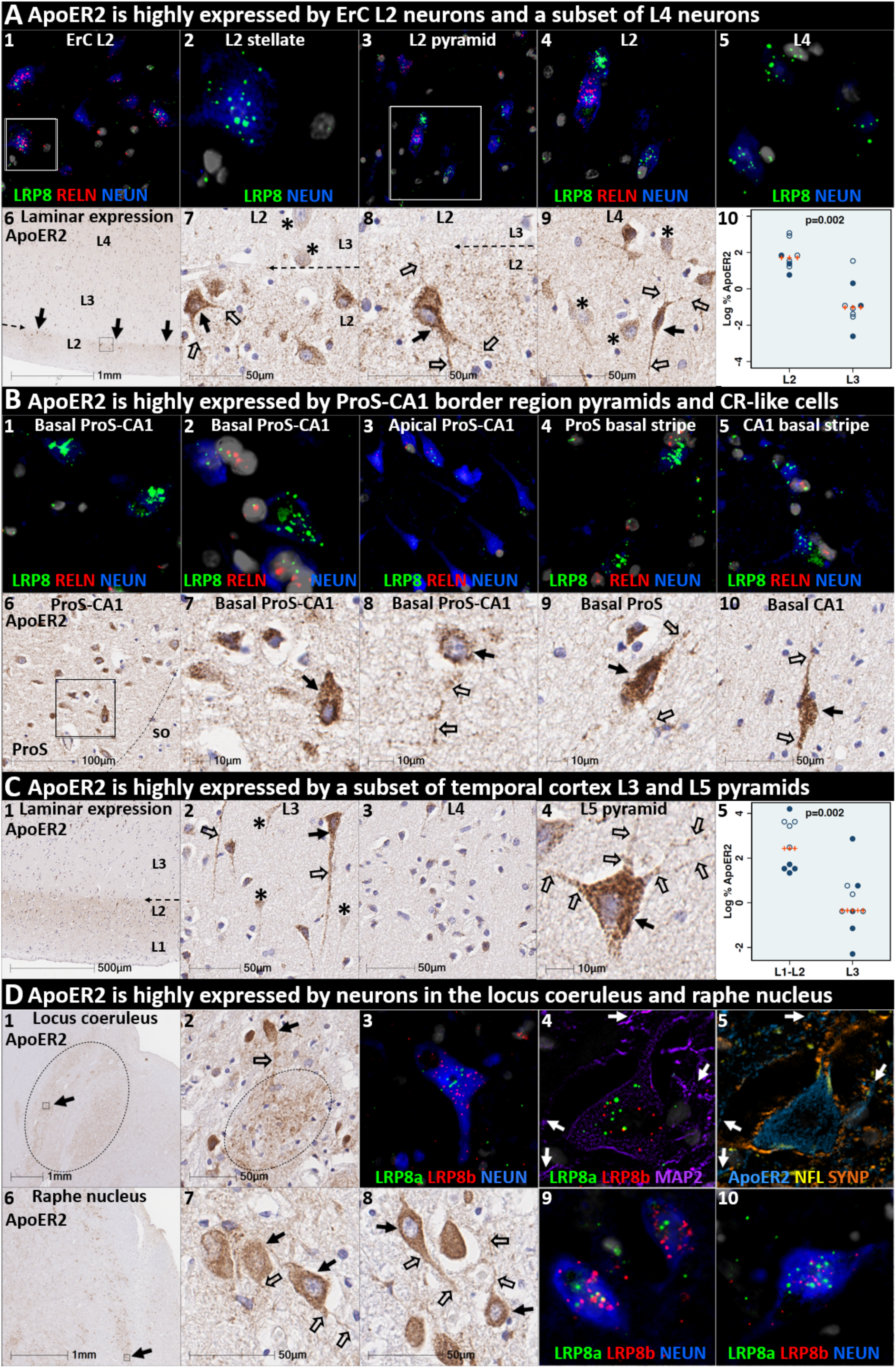
ApoER2 expression parallels regional, laminar, and cellular vulnerabilities to NFT pathology. Panels **A-D** are coronal sections of the ErC (**A**), ProS-CA1 border region (**B**), middle temporal gyrus (**C**), and transverse sections of the upper pons (**D**) from one representative middle-aged NFT stage I non-AD control. Soma and neuritic projections are demarcated by arrows and open arrows, respectively. **ErC** (**A_1-10_**): *LRP8* gene (**A_1-5_**) and ApoER2 protein (**A**_6-10_) are strongly expressed by stellate-shaped Reelin expressing neurons in (**A_1-2, 7-8_**), a subset of L2 pyramids (**A_3-4_**), and basal & apical dendritic tufts emanating from L2 neurons (**A_7-9_**). A visible laminar threshold (broken arrows in **A_6-8_**) for ApoER2 expression was observed near the L2-L3 border (**A_10_**). *LRP8* and ApoER2 are strongly expressed by a subset of L4 pyramids and surrounding neurites (**A_5_**, **A_9_**); adjacent L4 neurons with low ApoER2 expression are demarcated by an * in **A_9_**. **ProS-CA1 region** (**B_1-10_**): Strong *LRP8* (**B_1-5_**) and ApoER2 (**B_6-10_**) expression was observed in a subset of basal pyramids (**B_1-2_**) and neurons localized to the basal stripe (**B_4-5_**). Pyramids located in the middle and apical laminae tended to have lower *LRP8* and ApoER2 expression (**B_3_**). **Temporal neocortex** (**C_1-5_**): Strong ApoER2 expression in a subset of neocortical L3 and L5 pyramids, apical and basal dendritic projections (**C_3-4_**), and highly-ramified apical dendritic tufts (**C_1_**) located in the vicinity of L2. ApoER2 expression was low or absent in neighboring L3 pyramids (demarcated by * in **C_2_**) and most L4 stellate-shaped neurons (**C_3_**). A visible laminar threshold (broken arrow in **C_1_**) was observed near the L2-L3 border (**C_5_**). **Pontine LC & raphe nucleus** (**D_1-10_**): Strong ApoER2 (**D_1-2, 6-8_**) and *LRP8* (**D_3-5, 9-10_**) expression was observed in fusiform-shaped and multipolar LC neurons (**D_2-5_**) and in their MAP2-labeled dendritic tufts emanating into the peri-coeruleus region (oval in **D_2_**, white arrows in **D_4-5_**). ApoER2 (**D_6-8_**) and *LRP8* (**D_9-10_**) are also strongly expressed in neurons and neuritic projections in the raphe nucleus.

**Fig 4.**
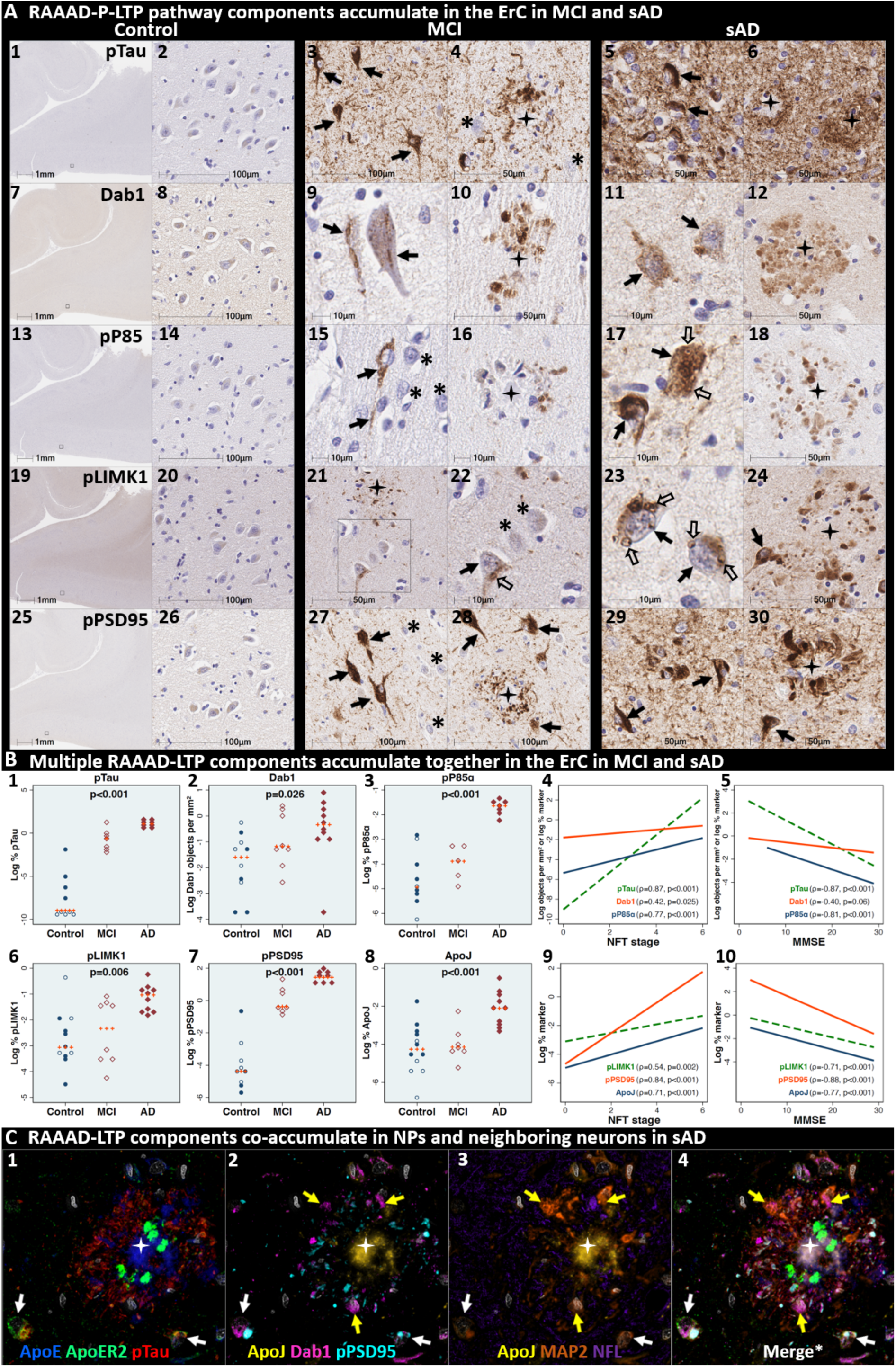
Multiple RAAAD-P-LTP pathway components accumulate together with pTau in ErC in MCI and sAD cases. (**A**) Serial coronal sections of the entorhinal region from representative non-AD control (NFT stage I), MCI (NFT stage III) and sAD (NFT stage VI) cases were probed with antibodies targeting RAAAD-P-LTP pathway components (see **Suppl Table 3**). In the non-sAD control (left column in **A**), IHC revealed very low expression of pTau, and little or no expression of pP85α_Tyr607_, pLIMK1_Thr508_, and pPSD95_Thr19_, with Dab1 expression confined to a subset of neurons. By contrast, in MCI (middle column in **A**) and especially sAD (right column in **A**), prominent accumulations of pTau, Dab1, pP85α_Tyr607_, pLIMK1_Thr508_, and pPSD95_Thr19_ were observed within abnormal neurons (solid arrows) and in the vicinity of NPs (black stars). Examples of neighboring neurons with little or no evidence of RAAAD-P-LTP component accumulation are designated with * (**A_4, 15, 22, 27_**). Open arrows in **A_17_** and **A_22-23_** designate granulovacuolar accumulations of pP85α_Tyr607_ and pLIMK1_Thr508_, respectively. (**B**) The expression of multiple neuronal RAAAD-P-LTP pathway components increased across the clinicopathological spectrum of sAD (**B_1-3, 6-8_**) and correlated with NFT stage (**B_4, 9_**) and cognitive deficits (**B_9, 10_**). Open and closed blue circles indicate young controls and age-matched controls; open and closed red diamonds indicate MCI cases and sAD cases, respectively. (**C**) MP-IHC revealed that Dab1, pTau, and pPSD95_Thr19_ accumulate together within ApoER2-expressing neurons (white arrows in **C_1-4_**) and that Dab1 accumulated within MAP2-labeled dystrophic dendrites (yellow arrows in **C_1-4_**) in the vicinity of ApoE and ApoJ-enriched extracellular plaques (white star).

## RESULTS

### ApoER2 is strongly expressed in the same regions, layers and neuron populations that develop NFT pathology in the earliest stages of sAD

ApoER2 regulates hippocampal dendritic arborization and memory in rodent models. ^39 60^ We recently demonstrated that human DG and hippocampal pyramidal neurons express ApoER2, ^46^ however, it is not yet known whether neurons that develop NFT pathology in the earliest stages of sAD express ApoER2. In the present study, IHC and MP-ISH/IHC revealed that regional, laminar, cellular and subcellular (dendritic) distributions of ApoER2/*LRP8* expression parallel well-established selective vulnerability to develop NFT pathology—with strong expression in ErC L2 neurons, the CA1-ProS border region basal stripe, a subset of L5 and L3 neocortical pyramids, and pontine LC and raphe nuclei neurons—and lower or absent expression in neuron populations that are spared from NFT pathology (**Fig 3, Ext Fig 3.1**). In ErC L2, both Reelin-expressing stellate projection neurons and pyramidal neurons are particularly vulnerable to pTau accumulation and are affected in NFT stage I,^2 3 8^ with pTau pathology originating in distal dendrites, followed by cell bodies and finally axons.^3^ L4 pyramids are affected next in NFT stage II, while L3 neurons are not affected until later in the disease process.^2 3 8^ In ErC L2, ISH and IHC in the ErC region revealed striking laminar and cellular *LRP8*/ApoER2 expression patterns, with strong signals observed within soma of Reelin-expressing stellate shaped neurons and a subset of pyramids lacking Reelin expression, and their basal and apical dendritic projections emanating into L1-L2 border region (**Fig 3, Ext Fig 3.1**). ApoER2 protein expression was higher in L2 than L3, resulting in a visible laminar termination threshold at the L2-L3 border. Strong ApoER2 expression was also observed in a subset of ErC L4 pyramids but was minimal or absent in a subset of neighboring L4 neurons (**Fig 3, Ext Fig 3.1**).

The basal stripe of the ProS-CA1 border region begins to accumulate pTau in NFT stage II and is the first subregion of the hippocampal formation to develop NFT pathology.^2 8^ ISH and IHC in the ProS-CA1 border region revealed prominent laminar and cell-specific *LRP8*/ApoER2 expression patterns with moderate to-strong expression in basal pyramids and non-pyramidal neurons and neurites localized to the basal stripe (**Fig 3**). Lower ApoER2 expression was observed within pyramids located within the middle and apical layers.

pTau accumulation is evident within multiple distal dendritic tips emanating from solitary temporal L5 and L3 pyramids very early in sAD (NFT stage I).^11^ The observation that pTau pathology spares neighboring neurons is an enduring puzzle that is not readily explained by prion-like Tau spread (see **Suppl Table 1**).^8 10 11^ Spiny stellate cells in L4 do not develop pTau pathology even in severe sAD.^11 61^ In temporal and frontal neocortex (**Fig 3, Ext Fig 3.1**), we observed striking layer and cell-specific distributions of ApoER2 with the strongest expression in the perikarya of subset of L5 and L2/L3 pyramids and their basal dendrites, and in the distal portions of their highly branched apical dendritic tufts in the L1-L2 border region (**Fig 3, Ext Fig 3.1**) ApoER2 protein expression was higher in L1-L2 than L3, resulting in a visible laminar termination threshold at the L2-L3 border. Despite this high expression in a subset of L5 neocortical pyramids and their apical dendrites (**Fig 3, Ext Fig 3.1**), ApoER2 expression was minimal or absent in a subset of neighboring L5 pyramids, and in most L4 neurons.

LC and raphe neurons accumulate pTau very early in the sAD process (NFT pre-tangle stages a/b and c, respectively).^9^ In the upper pons, IHC and MP-ISH/IHC revealed strong *LRP8*/ApoER2 expression in neurons and neuritic projections within LC and raphe nuclei (**Fig 3**), and scattered clusters of multipolar neurons located between both structures. Strong ApoER2 protein expression was observed within highly-branched projections emanating from LC neurons to the peri-coeruleus. MP-IHC revealed that ApoER2 expression in the LC had substantial overlap with MAP2-labeled dendritic arbors but minimal overlap with the axonal and pre-synaptic terminal markers NFL and synaptophysin, respectively.

The expression of two other ApoE receptors (LRP1 and VLDLR) was less restricted than ApoER2 and neither closely matched the laminar and cellular distribution of NFT pathology. LRP1 was expressed by glia and neurons, with prominent signals in glia surrounding NPs (**Ext Fig 3.2**). VLDLR was strongly and ubiquitously expressed by neurons in all neocortical layers including neocortical L4 stellate neurons (**Ext Fig 3.2**).

### Pathological accumulation of RAAAD-P-LTP pathway components in ErC L2

We next sought to determine if RAAAD-P-LTP pathway components accumulate in the ErC, and whether such accumulations increase across the clinicopathological spectrum of sAD. Single-target IHC using serial sections revealed that seven RAAAD-P-LTP pathway components—Dab1, pP85α_Tyr607_, pLIMK1_Thr508_, pTau, pPSD95_Thr19_, ApoJ and ApoE—accumulated in abnormal neurons and in proximity to NPs, were higher in MCI and sAD cases than controls, and positively correlated with histological progression or antemortem cognitive deficits (**Fig 4, Ext Fig 4.1**). RAAAD-P-LTP components exhibited different distributions and morphologies (**Fig 4A**). pTau prominently accumulated as hallmark NTs and NFTs, and in the neuritic components of NPs. Dab1 accumulated within intraneuronal inclusions and large globular complexes. pP85α_Tyr607_ and pLIMK1_Thr508_ accumulated in intraneuronal vacuolar structures reminiscent of GVDs and to a lesser extent in globular structures consistent with plaque-associated dystrophic neurites. pPSD95_Thr19_ accumulated in intraneuronal vacuolar structures, small punctae surrounding affected neurons, and globular structures consistent with plaque-associated dystrophic neurites. ApoE and ApoJ accumulated primarily within extracellular plaques. MP-IHC revealed that Dab1, pP85α_Tyr607_, pLIMK1_Thr508_, and pPSD95_Thr19_ generally accumulated together with pTau within MAP2 and ApoER2-labeled dystrophic dendrites and perikarya of nearby ApoER2-expressing abnormal neurons. Extracellular lipoprotein deposition was also evident with ApoE and ApoJ co-localized in the central core of many of the same NPs (**Fig 4C**).

We next searched for evidence for RAAAD-P-LTP pathway disruption in the earliest stage of sAD related pathology before clinical manifestations are apparent. Using IHC and MP-IHC for deep phenotyping of ErC in an NFT stage I non-AD case (**Fig 5**), we observed that Dab1, pLIMK1_Thr508_, and pPSD95_Thr19_ tend to accumulate together with pTau within the soma of the same L2 ApoER2-expressing stellate-shaped neurons and pyramidal neurons (**Fig 5**). Despite this selective vulnerability of their neighbors, a subset of adjacent neurons appears to be spared from pathological accumulation of pTau and other RAAAD-P-LTP components (designated with * in **Fig 5**). pTau and pPSD95_Thr19_ also accumulated within adjacent ApoER2-enriched neuritic projections. Dab1 accumulated within globular structures that co-localized with MAP2-labeled dystrophic dendrites, and to a lesser extent with NFL-labeled axons (**Ext Fig 5.1**). Unlike MCI and sAD cases, these intraneuronal and neuritic RAAAD-P-LTP pathway component accumulations were accompanied by only subtle ApoE and ApoJ deposits (**Ext Fig 4.1**), implying that intraneuronal RAAAD-P-LTP pathologies may precede overt extracellular lipoprotein deposition and plaque formation in the ErC.

**Fig 5.**
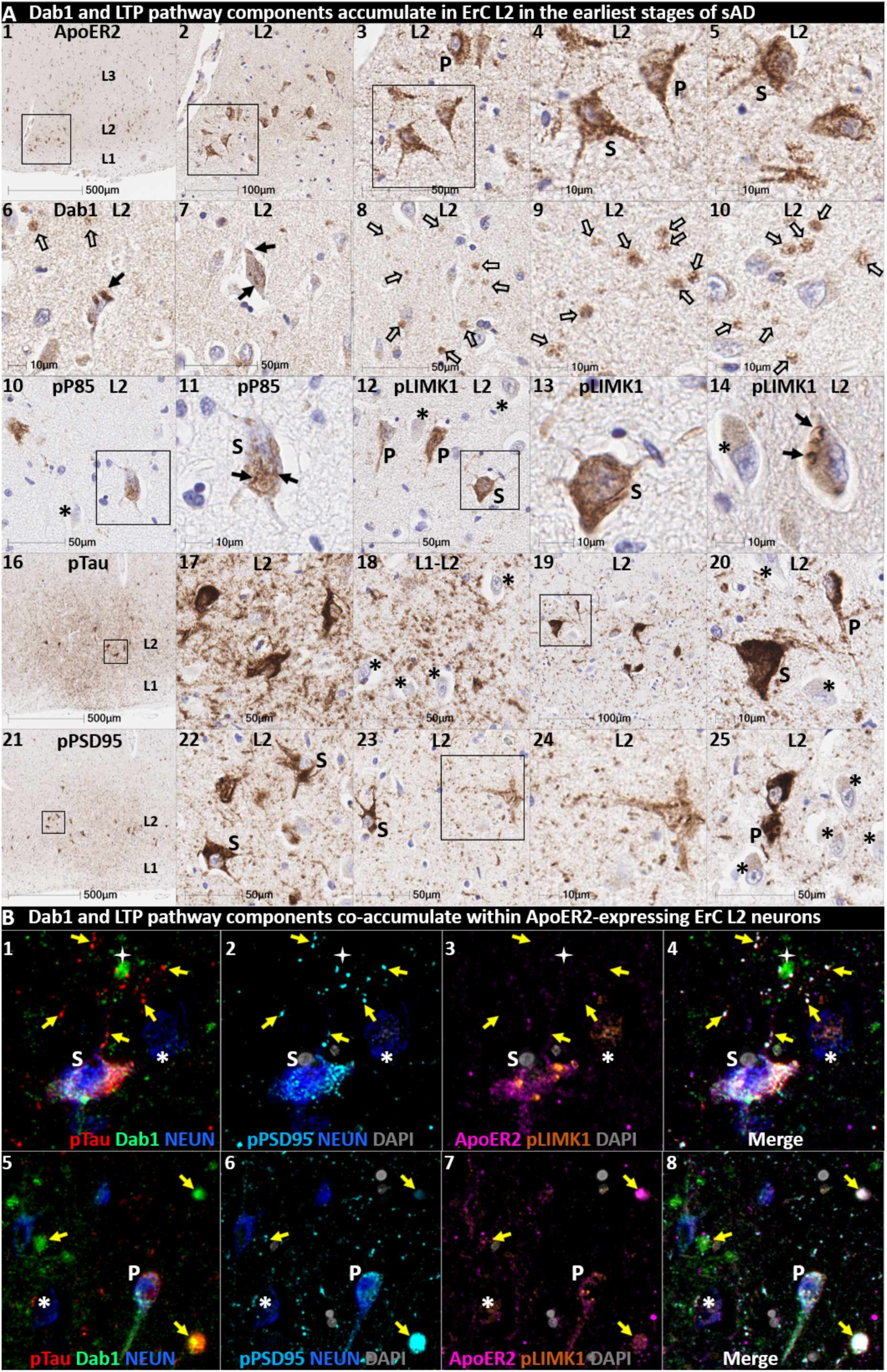
Multiple RAAAD-P-LTP components accumulate in the same ApoER2-expressing ErC L2 neurons in NFT stage I. Serial coronal sections of the ErC from one non-AD control case in the earliest stage of NFT pathology (NFT stage I, MMSE 27/30) were probed with antibodies targeting RAAAD-P-LTP pathway components. Single target IHC revealed that both stellate-shaped (designated S) and pyramid-shaped (designated P) neurons in ErC L2 strongly express ApoER2 (**A_1-5_**), and that multiple RAAAD-P-LTP pathway markers including Dab1 (**A_6-10_**), pP85α_Tyr607_ (**A_10-11_**), pLIMK1_Thr508_ (**A_12-15_**), and pPSD95_Thr19_ (**A_21-25_**) accumulated together with pTau (**A_16-20_**). Dab1 accumulated within the soma of a subset of ErC L2 neurons (solid arrows) and adjacent globular grape like structures (open arrows). pP85α_Tyr607_ accumulated within the soma of a subset of stellate-shaped neurons (S in **A_11_**). pLIMK1_Thr508_ accumulated within a subset of stellate-shaped (S in **A_12-13_**) and pyramidal neurons (P in **A_12_**). Granulovacuolar pLIMK1_Thr508_ accumulations are designated with solid arrows in **A_14_**. pTau accumulated within a subset of NFT-bearing stellate-shaped and pyramidal neurons, and as NTs in the adjacent neuropil. pPSD95Thr19 accumulated in the soma of affected stellate-shaped and pyramidal neurons and as discrete punctae in adjacent neuropil. Neighboring neurons with little or no evident accumulations of RAAAD-P-LTP components are designated with an * in **A_10-25_**. (**B**) MP-IHC revealed that pTau, Dab1, pLIMK1_Thr508_, and pPSD95_Thr19_ accumulated together within the same ApoER2-expressing stellate-shaped neurons (designated **S** in **B_1-5_**) and pyramidal neurons (designated **P** in **A_5-8_**), and adjacent dystrophic neurites (yellow arrows) that may emanate from these abnormal ApoER2 and Dab1 expressing ErC L2 neurons. Neighboring NEUN-labeled neurons with little or no RAAAD-P-LTP components evident in the soma are designated with an *.

### Pathological accumulation of RAAAD-P-LTP pathway components in the ProS-CA1 border region

We next sought to determine if RAAAD-P-LTP components accumulate in the ProS-CA1 border region, and whether such accumulations increase across the clinicopathological spectrum of sAD. Single-target IHC revealed that eight RAAAD-P-LTP pathway markers (Dab1, pP85α_Tyr607_, pLIMK1_Thr508_, pTau, pPSD95_Thr19_, pDab1_Tyr220_, ApoE, ApoJ) accumulated within abnormal neurons or within NPs, were higher in MCI and sAD cases than controls, and positively correlated with histological progression or cognitive deficits (**Figs 6-8, Ext Fig 6.1**). Peri-plaque Reelin aggregates were observed in ProS-CA1 in a subset of sAD cases (**Ext Fig 6.2**). However, these ProS-CA1 Reelin aggregates were less common and much less prominent than we previously observed in the CA2 region^46^ (**Ext Fig 6.2**), and did not correlate with histological progression or antemortem cognitive deficits (**Fig 6**). MP-IHC revealed that within the ProS-CA1 border region Dab1, pP85α_Tyr607_, and pPSD95_Thr19_ generally accumulated together with pTau in swollen dystrophic neurites surrounding ApoE enriched NPs, and within a subset of neighboring ApoER2-expressing neurons (**Figs 7-8**). In a recent publication ^46^, we described large plaque-associated Dab1 complexes in the molecular layer of the DG and the CA2 region of the hippocampus in sAD cases wherein Dab1 was primarily localized to MAP2-labeled dystrophic dendrites. In the present study, we observed similar large Dab1 complexes in three layers (SO, SP, SR) of ProS-CA1 border region in sAD cases (**Fig 7**). As expected, MP-IHC in the SP layer revealed that globular Dab1 primarily accumulated within MAP2-labeled swollen, dystrophic dendrites in the vicinity of ApoE-enriched NPs. Dab1-enriched dystrophic dendrites in the SP layer appear to emanate from one or more neighboring ApoER2-expressing neurons that co-accumulated multiple RAAAD-P-LTP components including pP85α_Tyr607_, pPSD95_Thr19_ and pTau (**Fig 7**). Unexpectedly, in the SR layer Dab1 accumulated primarily within NFL-positive (MAP2-negative) dystrophic axons surrounding ApoE-enriched NPs (**Fig 7**). In the SO layer, Dab1 accumulated in both MAP2-labeled dendrites and NFL-labeled axons (**Fig 7**), suggesting a dual dendritic and axonal origin for Dab1 in the vicinity of extracellular lipoprotein deposits.

**Fig 6.**
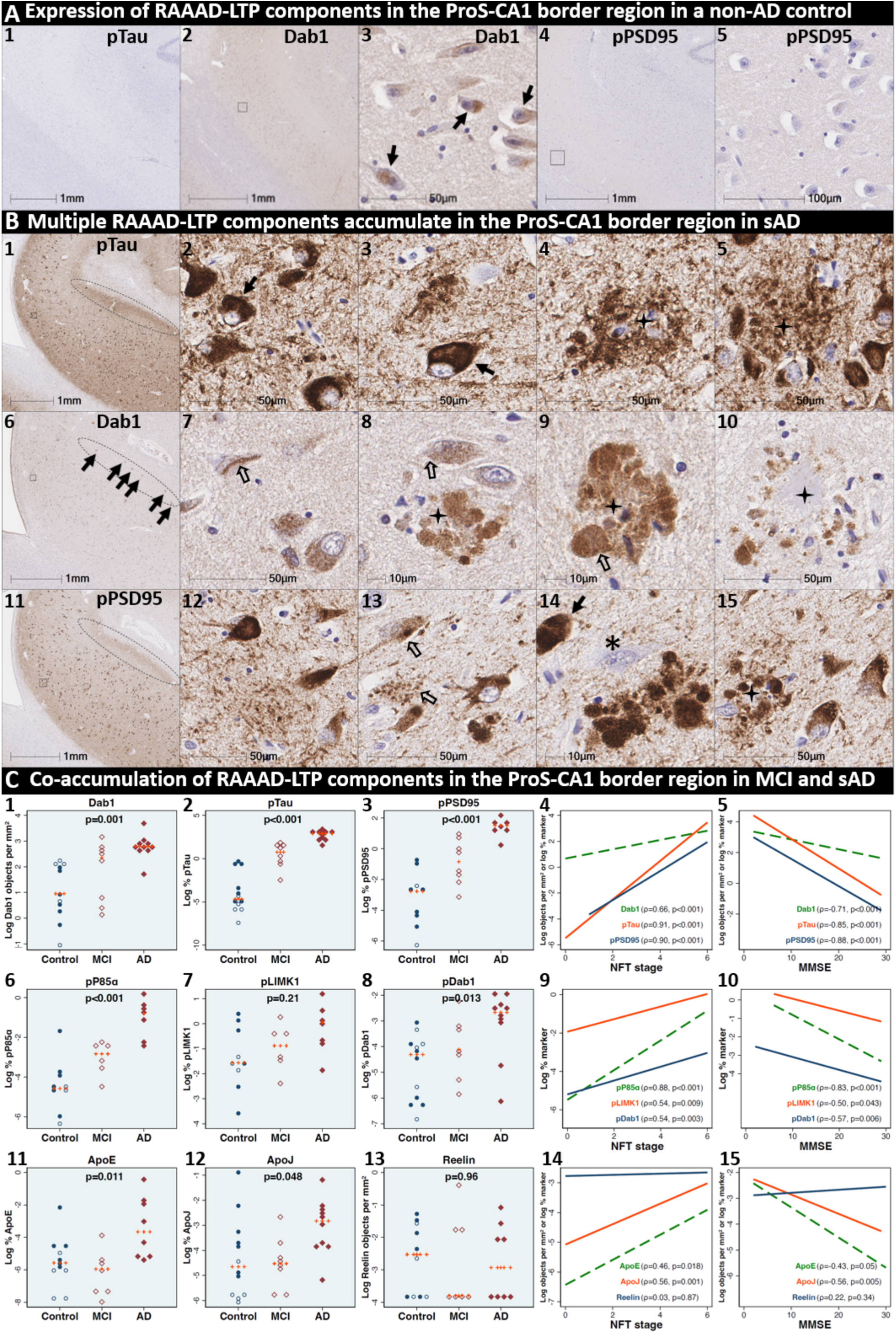
Co-accumulation of RAAAD-P-LTP pathway components in ProS-CA1 border region in sAD. (**A**) Serial coronal sections of the ProS-CA1 region from representative non-AD control (NFT stage I) and sAD (NFT stage IV) cases were probed with antibodies targeting RAAAD-P-LTP pathway components (see **Suppl Table 3**). In the non-sAD control (**A**), single-target IHC revealed little or no expression of pTau and pPSD95_Thr19_, with modest Dab1 expression confined to a subset of neurons. In sAD cases, pTau accumulated within NTs, NFTs, and the neuritic components of NPs (**B_1-5_**). Dab1 accumulated within abnormal neurons (**B_7-8_**) and in plaque-associated clusters of globular neurites (**B_8-10_**). pPSD95_Thr19_ accumulated within abnormal neurons, neuritic components of NPs, and in discrete punctae within the neuropil. All three components accumulated within the apical stripe region (arrows in **B_1, 6 &11_**), which harbors the terminal apical dendritic projections emanating from basal ProS-CA1 pyramidal neurons. (**C**) Expression of Dab1, pTau, pPSD95_Thr19_, pP85α_Tyr607_, pLIMK1_Thr508_, pDab1_Tyr220_, ApoE, and ApoJ increased across the clinicopathological spectrum of sAD and correlated with NFT stage and cognitive deficits (**C_1-15_**). Open and closed blue circles indicate young controls and age-matched controls; open and closed red diamonds indicate MCI cases and sAD cases, respectively. IHC images for pP85α_Tyr607_, pLIMK1_Thr508_, pDab1_Tyr220_, and ApoJ are provided in **Ext** Fig 6**.1**.

**Fig 7.**
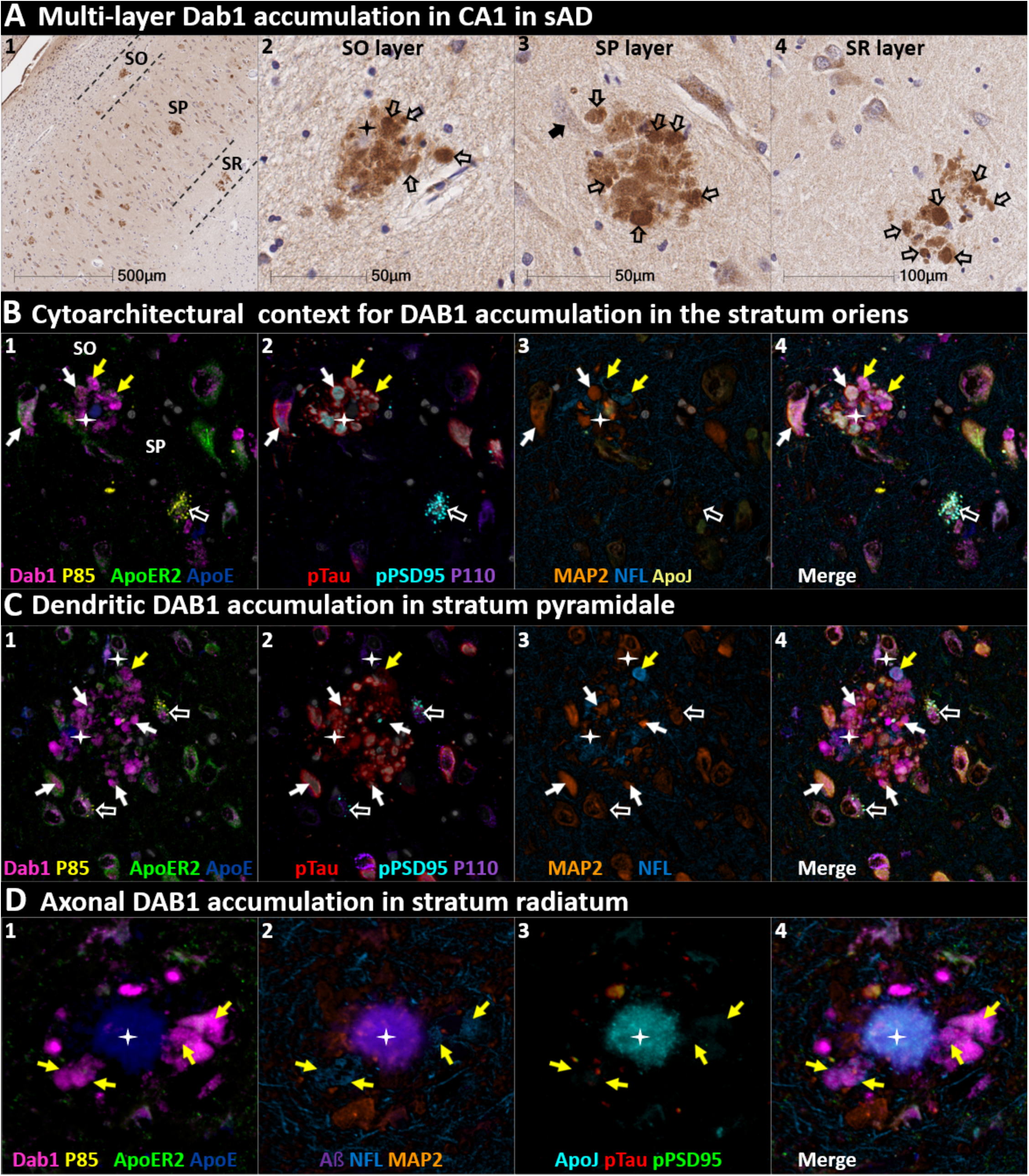
DAB1 accumulates in dystrophic dendrites and axons in CA1. (**A**) Single-target IHC revealed that Dab1 accumulates as grape-like clusters in three layers of CA1 (SO, SP, and SR). The globular Dab1 accumulations (open arrows in **A_2-4_**) are reminiscent of dystrophic neurites, however it is not yet clear if they are of dendritic or axonal origin. Dab1 clusters in the SP layer appear to emanate from a single neighboring neuron (black arrow in **A_3_**). A presumptive neighboring neuronal source of Dab1 complexes is not evident in the SO and SR. (**B**) MP-IHC revealed that in the SO layer Dab1 accumulates within both NFL-labeled axons (yellow arrows) and MAP2-labeled dendrites (white arrows) surrounding an ApoE enriched plaque (star). One neighboring ApoER2-expressing neuron at the SO-SP border had prominent intraneuronal accumulation of Dab1 and pTau (white arrows). Another nearby ApoER2 and Dab1-expressing neuron in the SP layer had prominent granulovacuolar co-accumulation of pP85α_Tyr607_ and pPSD95_Thr19_ (open white arrows), with little or no pTau evident. (**C**) MP-IHC in the SP layer revealed that Dab1 accumulates primarily within MAP2-labeled dendrites (white arrows in **C_1-4_**) and soma of neighboring neurons in the vicinity of extracellular ApoE (white stars). However, one Dab1-expressing globular structure co-localized with NFL (yellow arrow). Dab1 accumulated together with pTau and pPSD95_Thr19_ in several neighboring ApoER2-expressing neurons (white arrows in **C_1-4_**). Two nearby ApoER2 and Dab1-expressing neurons had prominent granulovacuolar co-accumulation of pP85α_Tyr607_ and pPSD95_Thr19_ (open white arrows), with little or no pTau evident. (**D**) MP-IHC in the SR layer revealed that Dab1 accumulated exclusively within dystrophic NFL-labeled axons (yellow arrows in **D_1-4_**) surrounding ApoE, ApoJ and Aβ-enriched extracellular plaques (white star). Unlike the SP layer, Dab1 accumulation was not evident within MAP2-labeled dendrites.

**Fig 8:**
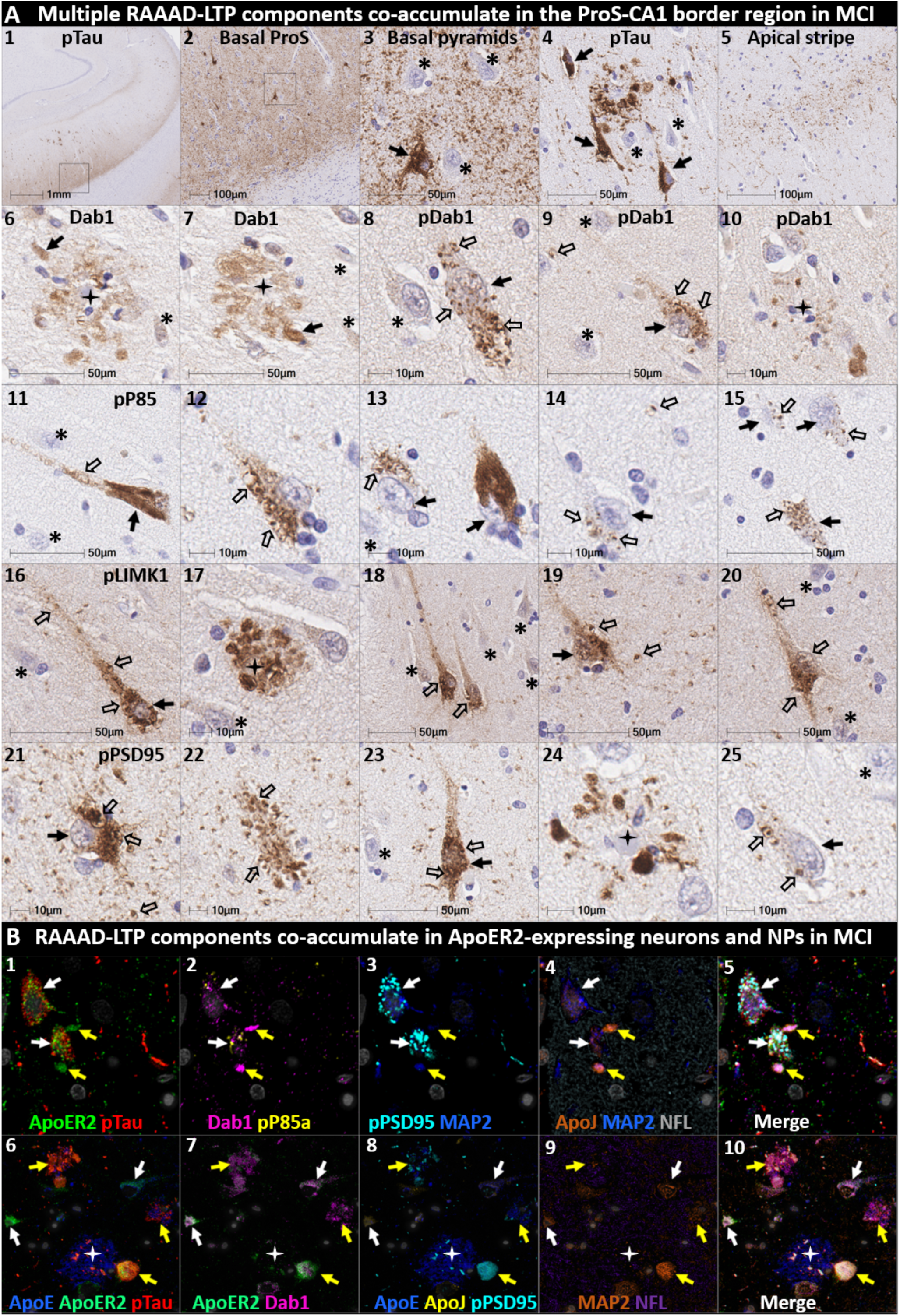
Multiple RAAAD-P-LTP components accumulate in ApoER2-expressing neurons and NPs in the ProS-CA1 border region in early MCI. Serial coronal sections of the ProS-CA1 border region in a representative early MCI case (NFT stage III, MMSE 28/30) were probed with antibodies targeting RAAAD-P-LTP pathway components. Single-target IHC revealed regional co-accumulation of multiple RAAAD-P-LTP pathway markers including pTau (**A_1-5_**), Dab1 (**A_6-7_**), pDab1 (**A_8-10_**), pP85α_Tyr607_ (**A_11-15_**), pLIMK1_Thr508_ (**A_16-20_**), and pPSD95_Thr19_ (**A_20-55_**). Affected neurons, GVD-like structures, and the apparent locations of extracellular plaques are designated with solid arrows, open arrows, and stars, respectively. Neighboring neurons with no evident accumulations of RAAAD-P-LTP components are designated with an * in **A_10-25_**. MP-IHC revealed that pTau, Dab1, pP85α_Tyr607_, and pPSD95_Thr19_ accumulated together within a subset of ApoER2-expressing basal pyramidal neurons (open arrows in **B_1-5_**), and within MAP2 and ApoER2 immunoreactive globular (yellow arrows in **B_1-10_**) or thread-like (white arrows in **B_1-5_**) dystrophic dendrites in the vicinity of affected neurons and ApoE-enriched NPs (white star in **B_6-10_**). NFL is not included in merged images (**B_5_**, **B_10_**) to enhance visualization.

We next sought to determine if accumulation of RAAAD-P-LTP components in the ProS-CA1 border region is evident in the early stages of sAD. Single-target IHC in an NFT stage III (MMSE 28/30) MCI case (**Fig 8**) revealed that even in this early stage when NFTs are sparse in the hippocampus^5^, pTau pathology was accompanied by prominent accumulations of Dab1, pDab1, pP85α_Tyr607_, pLIMK1_Thr508_, and pPSD95_Thr19_. Dab1 was most prominent within globular structures in the immediate vicinity of NPs. By contrast, pP85α_Tyr607_, pLIMK1_Thr508_ and pDab1 were most prominent in GVD-like vacuolar structures that were localized to the soma and neuritic projections, and as solitary vacuolar structures in the neuropil (open arrows in **Fig 8A**_4, 9&19_). pLIMK1_Thr508_ also labeled discrete punctae in neuropil (solid arrows in **Fig 8A**_6-10_). pPSD95 was prominent in GVD-like structures, small punctae surrounding affected neurons, and dystrophic neurites in the vicinity of NPs. MP-IHC revealed that Dab1, pP85α_Tyr607_, pTau and pPSD95_Thr19_ accumulated together within many of the same ApoER2-expressing abnormal neurons (**Fig 8C_1-5_**) and MAP2/ApoER2-labeled dystrophic dendrites in the vicinity of affected neurons and ApoE-enriched NPs (**Fig 8C_6-10_**).

### Dab1 accumulates together with RAAAD-P-LTP partners in temporal neocortex in MCI and sAD

Having shown that a subset of temporal L5 and L3 neocortical pyramids and their basal and apical dendritic projections strongly express ApoER2, we next sought to determine if RAAAD-P-LTP pathway components accumulate in the temporal neocortex and whether such accumulations increase across the clinicopathological spectrum of sAD. In a recent report using IHC in middle temporal gyrus sections ^46^, we observed that pPSD95_Thr19_ accumulates within abnormal neurons and NPs in MCI and sAD cases and correlates with NFT stage, Aβ plaque load, and cognitive deficits. In the present study, single-marker IHC using serial sections from the same cohort revealed that Dab1 accumulates in MCI and sAD cases (**Fig 9**) with strong signals localized to abnormal neurons, their basal dendritic arbors, and within the neuritic components of NPs. These Dab1-positive complexes increased across the spectrum of sAD and correlated with pPSD95_Thr19_, NFT stage, Aβ plaque load, and cognitive deficits (**Fig 9D**). MP-IHC revealed that Dab1 generally accumulated together with pTau and pPSD95_Thr19_ in many of the same ApoER2-expressing, NFT-bearing pyramids (**Fig 9E_1-5_**) and within swollen MAP2-positive dystrophic dendrites in the immediate vicinity of ApoE-enriched NPs (**Fig 9E_6-8_**). However, as previously shown in the ErC and ProS-CA1 border region, a subset of these peri-plaque Dab1 accumulations in the temporal neocortex co-localized with NFL-labeled dystrophic axons (**Fig 9E_10_**, **Ext Fig 9.1**).

**Fig 9.**
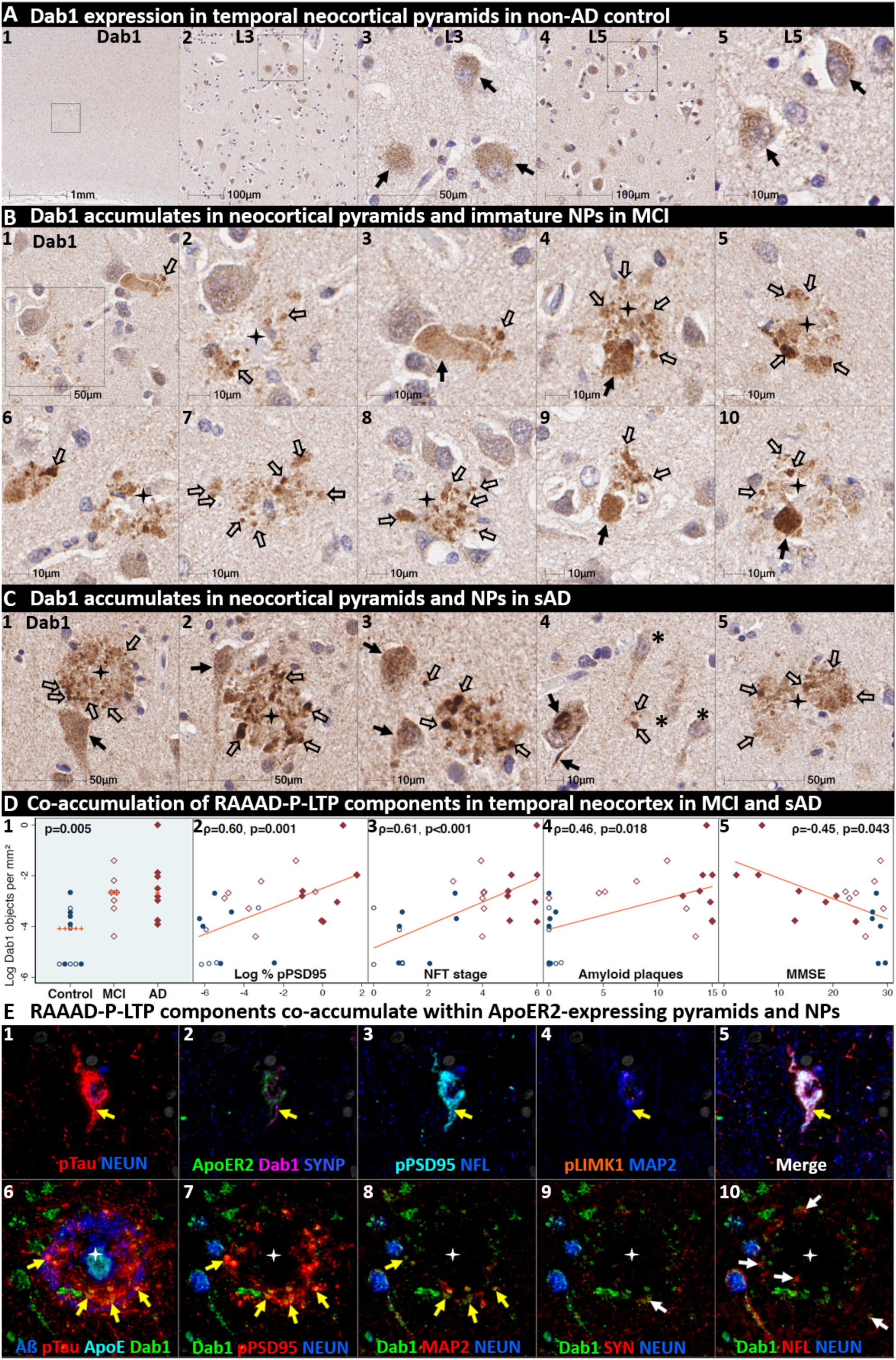
Dab1 accumulates together with RAAAD-P-LTP partners in temporal neocortex in MCI and sAD. (**A**) Serial coronal sections of the temporal neocortex from representative non-AD control (NFT stage I), MCI (NFT stage III) and sAD (NFT stage VI) cases were probed with antibodies targeting RAAAD-P-LTP pathway components (see **Suppl Table 3**). In the non-sAD control (**A**), single-target IHC revealed that Dab1 expression is confined to a subset of pyramidal neurons (solid arrows). In MCI (**B**) and sAD (**C**) cases, prominent accumulations of Dab1 were observed within abnormal neurons and within clusters of globular structures (open arrows in **B** and **C**) in the vicinity of NPs (black stars) that in some cases appeared to emanate from neighboring neurons (black arrows in **A_3-4_**, **A_9-10_**, **C_1-2_**). Intraneuronal Dab1 inclusions are evident in a subset of L3 and L5 pyramids (solid arrow **C4**); neighboring neurons with little or no Dab1 expression are designated with an * in **C_4_**. (**D**) Dab1 accumulation increased across the clinicopathological spectrum of sAD and correlated with pPSD95_Thr19_, NFT stage, Aβ plaque load, and cognitive deficits. Open and closed blue circles indicate young controls and age-matched controls; open and closed red diamonds indicate MCI cases and sAD cases, respectively. (**E**) MP-IHC revealed that Dab1, pTau and pPSD95_Thr19_ generally accumulated within the same ApoER2-expressing, NFT-bearing L5 and L3 pyramids (yellow arrows in **E_1-5_**) and MAP2-labeled dystrophic dendrites (yellow arrows in **E_6-10_**) in the immediate vicinity of Aβ plaques with ApoE enriched in the central core (white stars in **E_6-10_**). A small portion of Dab1 deposits appeared to accumulate within NFL-labeled axons or synaptophysin-labeled presynaptic terminals (white arrows in **E_9-10_**).

Since Aβ plaques can precede widespread pTau pathology in neocortex, we next searched for evidence of peri-plaque Dab1 accumulation in cases that have substantial neocortical Aβ plaques but minimal pTau pathology. Single-target IHC in a non-AD control with Aβ plaques (Thal phase 3) but no overt pTau pathology (NFT stage 0) revealed multiple, globular plaque-associated Dab1 accumulations that were most prominent in L5 and L3 (**Ext Fig 9.2**), with little or no pTau or pPSD95_Thr19_ evident in serial sections. MP-IHC revealed that some of these globular Dab1 accumulations were clustered around an ApoE-enriched central core (**Ext Fig 9.2**). When considered together with experimental evidence that ApoER2-Dab1 signaling regulates both conversion of AβPP to Aβ^62–64^ and GSK3β-mediated Tau phosphorylation ^40–44^, our findings suggest that Dab1 could potentially serve as a convergence point linking ApoE to both Aβ and pTau pathologies in early sAD. Future studies are needed to determine if these early plaque-associated Dab1 lesions are localized to axons or dendrites in temporal neocortex.

### Co-accumulation of RAAAD-P-LTP components in pontine LC-PC complex and raphe nucleus in sAD

We next sought to determine if Dab1 and other RAAAD-P-LTP pathway components accumulate in the vicinity of the LC and raphe nucleus in sAD cases and neurologically normal controls. In sAD cases, we observed that Dab1, pPSD95_Thr19_, and pTau accumulate in both the LC-PC complex and raphe nuclei, and in the vicinity of scattered clusters of multipolar neurons located between these two nuclei (**Fig 10** and **Ext Fig 10.1**). Accumulations of all three markers were less pronounced or absent in non-AD controls, with accumulations positively correlating with NFT stage (**Fig 10C**). Dab1 accumulated both as large globular neuritic complexes (**Fig 10B_2-10_**) and as intraneuronal inclusions (**Fig 10B_9_** & **Ext Fig 10.1**). Prominent pTau accumulations were observed in neuronal perikarya and in neighboring neuritic projections. pPSD95_Thr19_ accumulations had a similar distribution as pTau; however, unlike pTau, pPSD95_Thr19_ was most prominent within neuronal perikarya and discrete punctae surrounding affected neurons, with comparatively little staining of neuritic projections. MP-IHC revealed that Dab1 accumulated together with pPSD95_Thr19_ and pTau within many of the same ApoER2-expressing neurons and within MAP2-labeled dendritic arbors that appear to emanate from neighboring ApoER2-expressing neurons (**Fig 10D**). Extracellular accumulations of the ApoER2 ligands ApoE and ApoJ were observed in many sAD cases (**Ext Fig 10.2**). Reelin positive extracellular accumulations were observed in a subset of sAD cases (**Ext Fig 10.2**) but were not evident in most cases.

**Fig 10.**
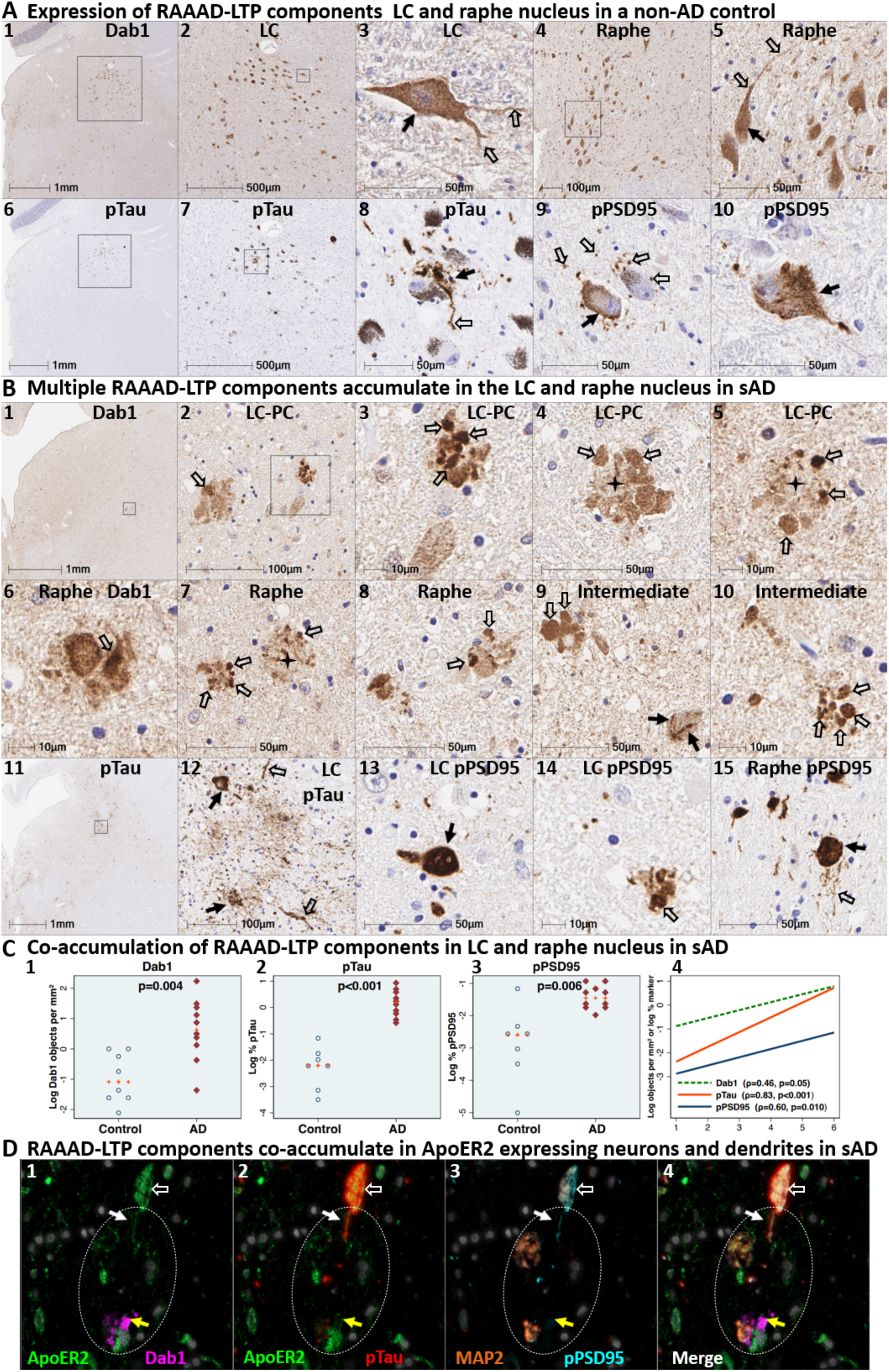
Dab1 accumulates together with RAAAD-P-LTP partners in LC and raphe nucleus in sAD. (**A**) Serial transverse sections of the upper pons from representative non-AD control (NFT stage I) and sAD (NFT stage V) cases were probed with antibodies targeting RAAAD-P-LTP pathway components (see **Suppl Table 3**). In the non-sAD controls (**A**), single-target IHC revealed that Dab1 is expressed by a subset of LC and raphe nucleus neurons (solid arrows in **A_3_, A_5_**) and their neuritic projections (open arrows in **A_3_, A_5_**). pTau and pPSD95_Thr19_ accumulated within small subsets of LC and raphe nucleus neurons (solid arrows in **A_8-10_**). pTau also accumulated in neuritic projections (open arrows in **A8**) while pPSD95 accumulated as discrete punctae in adjacent neuropil (open arrows in **A8**). In sAD cases (**B**), prominent accumulations of Dab1 were observed within clusters of globular structures (open arrows in **B_2-10_**) and as inclusions within abnormal neurons (solid arrows in **B_9_**). Additional examples of intraneuronal Dab1 inclusions are provided in **Ext** Fig 10**.1** (**C**) Dab1 accumulated across the clinicopathological spectrum of sAD and correlated with NFT stage. Open and closed blue circles indicate young controls and age-matched controls; open and closed red diamonds indicate MCI cases and sAD cases, respectively. (**D**) MP-IHC revealed that Dab1, pTau, and pPSD95_Thr19_ accumulated within the same ApoER2-expressing neurons (**D_1-4_**) and appeared to accumulate within MAP2-positive dendritic arbors (white circles in **E_1-4_**).

## Discussion

Fundamental questions about the origin and stereotypical progression of pTau pathology in sAD have remained unanswered for decades. Our data support a model in which ApoER2-Dab1 disruption is the underlying cause of pTau-related neurodegeneration in humans. In contrast to the prevailing hypothesis for NFT progression— that Tau spreads in a prion-like manner—our findings suggest that pTau is locally produced by ApoER2 expressing neurons at multiple anatomical locations in response to ApoER2-Dab1 disruption. Since the RAAAD-P-LTP pathway regulates memory through delivery of essential lipids and stabilization of actin, microtubules, and synapses (**Fig 2B**), our finding that multiple pathway components co-accumulate in each affected region reframes Tau pathology as one of many consequences of pathway disruption. As an alternative to prion-like spread, ApoER2-Dab1 disruption provides a single, shared mechanism that can: (1) explain the actin and microtubule destabilization, synaptic dysfunction, extracellular lipoprotein deposition, and cognitive deficits that characterize sAD; (2) mechanistically link the etiology of NPs to NTs, NFTs and GVDs; and (3) integrate hallmark ApoE, pTau, and Aβ pathologies into a unifying model for sAD in humans.

### Can ApoER2 expression and demand for activation explain the origin(s) and progression of NFT pathology?

ApoER2-Dab1 disruption promotes Tau hyperphosphorylation through compromised Reelin signaling. ^39–43^ Since ApoER2 is not ubiquitously expressed, our finding that ApoER2 is strongly expressed in the same regions, layers, neurons, and subcellular compartment (distal dendritic tips) ^8 11 65^ that accumulate pTau early in sAD is consistent with our model in which multisite ApoER2 disruption drives development of NFTs (**Fig 1**). Whereas the presence of ApoER2 may be required for NFT formation, a high demand for RAAAD-P-LTP pathway activation and turnover could also contribute to the selective vulnerability of ApoER2-expressing neurons. It is therefore notable that the ErC and LC are active nearly continuously from birth until death, due to their pivotal roles in memory formation during waking hours and memory consolidation during sleep. ^68–71^

### Dab1 accumulation reveals evidence for Reelin-ApoER2-Dab1 pathway disruption

Activation of the Reelin-ApoER2-Dab1 cascade shapes and strengthens synaptic connections underlying learning and memory (**Fig 2B**).^34 37 44 66–69^ Since Reelin signaling through ApoER2 induces rapid proteasomal degradation of Dab1, ^70 71^ Dab1 protein accumulation implies a localized, functional deficit in Reelin signaling through ApoER2.^46^ Our recent finding that Dab1 accumulates within dystrophic dendrites in the terminal zones of the perforant path in sAD ^46^ provided evidence for ApoER2-Dab1 disruption in neuronal circuitry underlying memory in humans. In the present study, our finding that Dab1 accumulates in each of five sampled regions that develop NFT pathology prior to the hippocampus and DG provides evidence that ApoER2-Dab1 disruption is widespread even in the early stages of sAD. Remarkably, Dab1 accumulation was extensive in MCI, and even preceded overt pTau accumulation in some controls, indicating that ApoER2-Dab1 disruption is likely a very early degenerative phenomenon. MP-IHC revealed that Dab1 and pTau generally accumulated within the same layers, neurons, and subcellular compartment (MAP2-labeled dystrophic dendrites) in each region. Since Dab1 is an upstream regulator of GSK3β-mediated Tau phosphorylation (**Fig 2B**),^40–44^ together these observations are consistent with our model wherein the accumulations of Dab1 and pTau (in NTs, NFTs, NPs) in each region result from RAAAD-P-LTP pathway disruption in ApoER2-expressing neurons. Intriguingly, Bracher-Smith et al.^72^ recently reported a genetic association between the *DAB1* locus and AD risk that was evident only in *APOE4* homozygotes, a high-risk population that accounts ≈10% of sAD cases.^73^ Our findings of extensive Dab1 accumulation in *APOE3* homozygote and *APOE2/APOE3* heterozygote MCI and sAD cases provide evidence that ApoER2-Dab1 disruption is a shared mechanism underlying sAD that may be exacerbated by, but is not dependent on, the *APOE4* gene variant. Moreover, our findings that Dab1 accumulates together with phosphorylated forms of P85α and three downstream ApoER2-Dab1-P85α/PI3K signaling partners (LIMK1, Tau, PSD95) that stabilize actin, microtubules, and postsynaptic complexes, respectively, (**Fig 2B**) ^46 74–76^ provide insights into molecular pathways and mechanisms linking Dab1 accumulation to cytoskeletal instability and synapse loss in sAD.

### Dendritic co-accumulation of pP85α, pLIMK1, pTau and pPSD95 as evidence for ApoER2-Dab1 disruption

Dendritic spines are dynamic actin-rich protrusions that harbor excitatory synapses within postsynaptic densities, whose plasticity plays a central role in learning and memory.^77 78^ Binding of Reelin to ApoER2 stabilizes actin and microtubule cytoskeletons and postsynaptic densities by regulating phosphorylation and activation of Dab1, PI3K, LIMK1, GSK3β, Tau and PSD95 (reviewed in ^46^ (**Fig 2B**)).^34 41–44 66 68 69 74–76 79–81^ In rodent and cellular models, compromised Reelin-ApoER2-Dab1-PI3K signaling promotes GSK3β-mediated Tau hyperphosphorylation and somatodendritic localization.^40–44^ The Dab1 PTB domain—which recruits ApoER2-Dab1 signaling complexes to lipid rafts by simultaneously binding the polar head group of PIP2 and cytoplasmic tail of ApoER2 ^82^(**Fig 2B**)—is required for Reelin-Dab1 signaling.^83 84^ P85α serves as a critical node in this pathway by recruiting Dab1 and PI3K to lipid rafts,^85 86^ enabling formation of postsynaptic ApoER2-Dab1-PI3K-signaling complexes (**Fig 2B**). Reelin triggers interactions between P85α and Dab1 to suppress GSK3β-mediated Tau phosphorylation via activation of the PI3K pathway,^86^ while Tyr607 phosphorylation of P85α conversely inhibits PI3K activity.^87^ Since activated GSK3β phosphorylates PSD95 to induce synapse disassembly,^81^ Reelin-ApoER2-Dab1 pathway disruption provides a straightforward mechanism that could account for somatodendritic accumulations of both pTau and pPSD95_Thr19_ (reviewed in ^46^). LIMK1 phosphorylation regulates remodeling of the actin cytoskeleton of dendritic spines.^88 89^ Heredia et al.^90^ observed an increase in number of pLIMK1_Thr508_ positive neurons in AD-affected regions. Similarly, in human hippocampus in sAD, we previously reported that pLIMK1_Thr508_ and pP85α_Tyr607_ accumulate together within GVD-like structures in neurons that accumulate pTau and pPSD95_Thr19_, and in neighboring abnormal neurons lacking overt pTau pathology.^46^ In the present study, we observed that five core intraneuronal pathway components (Dab1, pP85α_Tyr607_, pLIMK1_Thr508_, pTau, PSD95_Thr19_) accumulate together within many of the same ApoER2-expressing, NFT and/or GVD-bearing neurons and MAP2-labeled dystrophic dendrites in each region. These collective observations provide multifaceted evidence for a pathogenic nexus centered around dendritic ApoER2-Dab1 disruption in the neuron populations that are most vulnerable to early NFT pathology.

### ApoER2-Dab1 disruption can explain pTau pathology without invoking prion-like spread

The Tau prion-like connectome-based spread model has several discrepancies that are not easily reconciled with spatiotemporal distributions of pTau pathology in human brain (see **Suppl Table 1** for detailed description). Our finding that multiple RAAAD-P-LTP pathway components, including several that are upstream of Tau phosphorylation (**Fig 2B**), accumulate in each affected region strongly suggests that pTau is locally produced by ApoER2-expressing neurons at each site. Since prion-like properties are thought to be a unique feature of Tau, ^91 92^ connectome-based spread of numerous RAAAD-P-LTP components is an exceedingly unlikely explanation for this multisite co-accumulation. Tau inclusions are known to originate in distal dendrites. ^8 11 65^ Thus, our finding that pTau accumulates together with multiple upstream and downstream RAAAD-P-LTP components in MAP2-labeled dystrophic dendrites of ApoER2-expressing neurons—revealed by advanced MP-IHC methods—is consistent with ApoER2-Dab1 disruption but not easily reconciled with prion-like spread. The prion-like spread hypothesis, as usually presented, also does not explain the point(s) of origin of the disease process, or which intrinsic molecular features account for the vulnerability of this origin.^16^ By contrast, dendritic ApoER2-Dab1 pathway disruption could explain the local production and accumulation of pTau in all affected neurons including the neuron populations where pTau pathology is classically reported to begin (LC, ErC L2).

### Dendritic ApoER2-Dab1 disruption can mechanistically & spatially link four pTau-containing lesions

Dendritic ApoER2-Dab1 disruption provides a plausible shared mechanism that can link four hallmark pTau containing pathologies (NTs, NFTs, NPs, GVDs) (**Fig 2C**). We previously reported that two ApoER2 ligands (ApoE and Reelin) accumulate together with extracellular Aβ and dendritic pTau in hippocampal NPs.^46^ Our present finding that yet another ApoER2 ligand (ApoJ)^45^ accumulates together with ApoE in the central core of many NPs, provides further evidence for ApoE receptor-ligand disruption and extracellular lipoprotein deposition in plaque-associated dystrophic neurites. ApoER2, Dab1, P85α and PSD95 are enriched in the distal dendritic tips ^35 68 93–95^ of pyramidal neurons, where pTau inclusions originate as NTs before progressing to proximal dendrites and soma of NFT-bearing neurons.^8 11^ Taken together, these findings support a model wherein dendritic ApoER2-Dab1 disruption initiates a disease-cascade resulting in both ‘extracellular trapping’ of ApoER2 ligands with lipoprotein deposition in the synaptic cleft, and accumulation of ApoER2-Dab1 signaling partners in adjacent distal dendrites (reviewed in ^46^)(**Fig 2C**). GVDs are enigmatic pTau-containing lesions that are first evident in hippocampal and subicular pyramids that are considered to be pre-NFTs by some investigators.^96–99^ Nishikawa et al ^100 101^ showed strong expression of PIP2 and lipid raft proteins in GVDs and NFTs, suggesting that both lesions originate in PI-enriched lipid rafts. Our finding that phosphorylated forms of RAAAD-P-LTP components known to play central roles in organizing lipid rafts—including P85α, Dab1, PSD95, and LIMK ^68 93–95^—accumulate alongside pTau in GVDs provides a mechanistic link between ApoER2 Dab1 pathway disruption, granulovacuolar degeneration and NFT formation. Thus, dendritic ApoER2 disruption is plausible mechanism that could help explain why NPs, NTs, NFTs and GVDs develop within spatially separated substructures (synaptic cleft, dendritic tips, soma) in the same affected neurons (**Fig 2C**). Consistent with this interpretation, in the ProS-CA1 border region we observed that multiple RAAAD-P-LTP pathway components accumulate in anatomically distinct areas (basal stripe, pyramidal layer, apical stripe) corresponding to spatially-separated substructures in the same neuron populations.

### Is ApoER2-Dab1 disruption also the origin of Aβ pathology?

Dab1 serves as a cytoplasmic adaptor protein for both ApoER2 and AβPP.^102 103^ ApoER2-Dab1 signaling has been shown to regulate AβPP cleavage and Aβ synthesis in model systems ^62–64^. AβPP is localized to axons,^104, 105^ and Aβ appears to be synthesized primarily in dystrophic axon terminals surrounding Aβ plaques.^106 107^ Although they are most abundant in dendritic spines, emerging evidence indicates that ApoER2 and Dab1 are also enriched in axonal growth cones,^42^ where they regulate axonal arborization in a Reelin and ApoE dependent manner.^108–110^ Thus, our finding that Dab1 accumulates in a subset of NFL-labeled dystrophic axons surrounding Aβ plaques suggests that axonal ApoER2-Dab1 pathway disruption may regulate Aβ formation in humans. Together with experimental evidence that ApoER2-Dab1 signaling regulates GSK3β-mediated Tau phosphorylation ^40–44^ and our findings that Dab1 accumulates together with pTau in dystrophic dendrites, our findings suggest that ApoER2-Dab1 disruption and Dab1 accumulation could serve as a convergence point and biologically plausible shared origin linking ApoE to the Aβ plaques and pTau tangles that define sAD in humans.^111^

### RAAAD-P-LTP disruption integrates formerly disjointed observations into a unifying model for sAD

There is currently no hypothesis that can integrate ApoE with hallmark pTau and Aβ pathologies, selective vulnerability of entorhinal and pontine substructures, and genetic risk factors to satisfactorily explain sAD pathogenesis in humans. Our present and previous^46^ findings provide the foundation for a unifying model wherein ApoE receptor-ligand disruption triggers a disease-cascade that ultimately manifests as sAD by: (1) disrupting ApoE receptor-ApoE/ApoJ dependent delivery of lipid cargo required to shape and remodel neuronal membranes; (2) disrupting RAAAD-P-LTP signaling cascades that stabilize actin and microtubule cytoskeletons and postsynaptic receptor complexes; (3) promoting Tau hyperphosphorylation and NT/NFT formation; (4) trapping lipid-laden ApoE/ApoJ particles outside neurons where they provide seeds for Aβ oligomerization;^112–114^ and (5) promoting axonal Aβ synthesis and Aβ plaque formation. This model is attractive because it can help explain the origins of pTau pathology in the LC, ErC L2 and dendritic tips of solitary L5 and L3 neocortical pyramids and the sequential involvement of other neuron populations in successive NFT stages (**Fig 1**, **Suppl Table 1**).^11^ The model also provides a straightforward, plausible explanation for the convergence of Aβ with pTau, ApoE, ApoJ and other RAAAD-P-LTP pathway components in NP niche, and mechanistically links the etiology of NPs to NTs, NFTs and GVDs. The RAAAD-P-LTP pathway disruption model is also consistent with established and emerging genetic risk factors for sAD ^115 116^—including variants in genes coding for ApoE,^115 117–120^ ApoJ,^116 121^ Dab1,^72^ P85α,^122 123^ glial pathways that govern extra-neuronal clearance of ApoE, ApoJ, Aβ, and lipids ^115 124 125^ and neuronal pathways that govern proteasomal and endolysosomal function ^116 126^.

### Implications for sAD therapeutics

The prion-like spread model provides the rationale for immunotherapeutics designed to block the spread of Tau throughout the brain.^18 19^ However, despite clear beneficial effects in mice that are genetically modified to overexpress Tau,^127 128^ anti-Tau monoclonals tested thus far in MCI and early sAD patients failed to demonstrate efficacy or even worsened cognitive decline.^129 130^ Since our findings suggest that pTau accumulation in humans results from multisite ApoER2-Dab1 disruption rather than pTau spread, drugs targeting underlying causes of this disruption may be better positioned to delay sAD progression.

### Underlying causes of ApoER2-Dab1 pathway disruption

Plausible triggers for ApoER2-Dab1 pathway disruption include lipid peroxidation, injury-induced ApoE hypersecretion, and Reelin depletion. ApoE particles deliver lipids that are highly vulnerable to peroxidation to neuronal ApoE receptors including ApoER2. ^46^ Our recent finding that ApoE and ApoER2 are vulnerable to lipid aldehyde-induced adduction and crosslinking ^46^ provides a mechanistic link between lipid peroxidation and ApoER2-Dab1 disruption. Since lipid peroxidation is a common feature of many sAD risk factors (reviewed in ^46^) this mechanism could help explain previous observations that lipid peroxidation is markedly increased in the early stages of sAD. ^46 131–140^ Astrocytes stimulate repair of injured axons by dramatically increasing local secretion of lipid-loaded ApoE particles ^141 142^. Since ApoE competes with Reelin for ApoER2 binding in model systems,^143^ an excess of ApoE following injury could potentially compromise Reelin ApoER2-Dab1 signaling.^143^ Lastly, since Reelin-secreting neurons are reported to degenerate in sAD, ^144–147^ Reelin depletion could help explain (or exacerbate) the hyperphosphorylation of Tau ^40–44^ as well as the accumulations of Dab1, pP85α_Tyr607_, pLIMK1_Thr508_, and pPSD95_Thr19_ observed in the present study. However, unlike ApoE receptor-ligand disruption, Reelin depletion alone cannot readily account for our finding that three ApoER2 ligands (ApoE, ApoJ, Reelin) accumulate in the extracellular space in the immediate vicinity of many NPs.

### Strengths and limitations

Strengths of the present study include: (1) use of rapidly-autopsied specimens that underwent uniform, time limited (48h) fixation,^81 148 149^ in conjunction with antemortem cognitive data; (2) use of orthogonal methods including single-marker IHC with multi-epitope labeling using several independent antibodies and RNA-protein co-detection; and (3) pathological and cytoarchitectural context provided by MP-IHC and MP-ISH/IHC. Human brain IHC studies with autopsy delay of up to 72 hours and fixation for months to years are routinely published. Delayed autopsy and prolonged fixation—which requires harsh conditions for antigen retrieval—are reported to have limited impact on highly-aggregated proteins such as Aβ and pTau ^148^ but can obscure signals for less aggregated proteins.^149–151^ Thus, use of rapidly-autopsied, and uniformly and minimally-fixed brain specimens is an important strength of the present study. Postmortem specimens spanning the clinicopathological spectrum of sAD were used to approximate the spatiotemporal sequence of NFT progression. This cross-sectional study design cannot establish a sequence of disease progression.^152^ The moderate sample sizes (n=64 for most markers) are an important limitation. Although RAAAD-P-LTP pathologies were observed in all major *APOE* variants and both sexes, larger studies are needed to determine if results are influenced by genetics, sex, and other variables. The comparable IHC results obtained using specimens from three brain banks that employed different procedures for autopsies, sample processing and pathological assessment add confidence to study findings. However, since all three cohorts were primarily of Caucasian descent, future studies including more races and ethnicities are needed to determine generalizability. The human brain contains multiple *LRP8* splice isoforms ^153 154^ that impact ligand-binding, receptor complex formation, synaptic plasticity and memory.^33 35 154–, 157^ The present study used two mRNA probes designed to detect most (but not all) *LRP8* isoforms. Future studies are therefore needed to characterize distributions of *LRP8* splice variants. Although they lack the restricted distribution of ApoER2, two other brain ApoE receptors—LRP1 and VLDLR—that share ligands and adaptor proteins with ApoER2 may contribute to observed disease manifestations via overlapping mechanisms. While our finding that pTau accumulated together with upstream markers of pTau production (i.e., Dab1, pP85α_Tyr607_) strongly suggests that pTau is locally produced by neurons within each affected region, it does not rule out the possibility that prion-like propagation of Tau—including pTau or Tau that is not detected by traditional methods ^158–160^ —could contribute to pTau-related neurodegeneration.

### Summary & Conclusion

We found that five neuron populations that accumulate pTau in the earliest stages of sAD strongly express ApoER2 and that pTau is but one of numerous RAAAD-P-LTP components that accumulate within NPs and NT-, NFT-, and GVD-bearing ApoER2-expressing neurons. Collective findings provide the basis for the RAAAD-P-LTP hypothesis, a unifying model that implicates dendritic ApoER2-Dab1 disruption at multiple anatomical locations as the major driver of both Tau hyperphosphorylation and neurodegeneration in sAD. This model provides a new conceptual framework to explain why specific neurons degenerate in sAD, mechanistically & spatially links four pTau-containing lesions, integrates hallmark ApoE, pTau and Aβ pathologies into a unifying model, and identifies RAAAD-P-LTP pathway components as potential mechanism based biomarkers and therapeutic targets for sAD in humans.

## Declaration of competing interest

The National Institutes of Health has filed patent applications related to mechanism-based biomarkers that are broadly related to this manuscript, with one coauthor (CER) named as an inventor.

## Data Availability

The datasets and code are available from the corresponding author upon reasonable request.

## Abbreviations

AD: Alzheimer’s disease
ApoE: Apolipoprotein E
ApoJ: Apolipoprotein J
ApoER2: ApoE receptor 2
CA: Cornu ammonis
CR: Cajal-Retzius
Dab1: Disabled homolog-1
DG: Dentate gyrus
ErC: Entorhinal region
GVD: Granulovacuolar degeneration body
L2: Layer II
LC: Locus coeruleus
LIMK1: LIM domain kinase-1
*LRP8*: Low-density lipoprotein receptor-related protein 8 (gene encoding ApoER2)
LTP: Long-term potentiation
NEUN: neuronal nuclear/soma antigen
NFL: Neurofilament light chain
NFT: Neurofibrillary tangle
NP: Neuritic plaque
NT: Neuropil thread
P85α: PI3K regulatory subunit P85alpha
PC: Peri-coeruleus
PI: Phosphatidylinositol
PIP2: Phosphatidylinositol-(4,5)-bisphosphate
PI3K: Phosphatidylinositol-3-kinase
PMI: Postmortem interval
ProS: Prosubiculum
PSD95: Postsynaptic density protein 95
pTau: pTau_Ser202/Thr205_
RAAAD-P-LTP: Reelin/ApoE/ApoJ-ApoER2-Dab1-P85-LIMK1-Tau-PSD95
*RELN*: gene encoding Reelin protein
sAD: sporadic AD
SO: Stratum oriens
SP: Stratum pyramidale SR Stratum radiatum
SYNP: Synaptophysin

## Acknowledgements

This project was supported by the intramural programs of NIA, NIAAA and NINDS, NIH. Additional support was provided as a research gift from John M. Davis to the Laboratory of Clinical Investigation, NIA/NIH. The MP-IHC work utilized the computational resources of the NIH HPC Biowulf cluster (http://hpc.nih.gov). We are grateful to the research volunteers and the UA-HBB, BBDP, and UK-ADRC research teams including Henry Waldvogel, Clinton Turner and Marika Ezses (UA-HBB) and Geidy Serrano (BBDP). The UA-HBB is supported by the Neurological Foundation of New Zealand. The BBDP is supported by the NINDS (U24 NS072026 National Brain and Tissue Resource for Parkinson’s Disease and Related Disorders), the NIA (P30 AG19610 Arizona Alzheimer’s Disease Core Center), the Arizona Department of Health Services (contract 211002, Arizona Alzheimer’s Research Center), the Arizona Biomedical Research Commission (contracts 4001, 0011, 05-901 and 1001 to the Arizona Parkinson’s Disease Consortium) and the Michael J. Fox Foundation for Parkinson’s Research. The UK-ADRC is supported by the NIA/NIH (P30 AG072946).

We thank Pragun Vohra and the Histoserv staff for immunohistochemical expertise and immunostaining. We are grateful to Elizabeth Calzada (NIA) for editing the manuscript and Marc Raley (Visual Media Services, NIA/NIDA) for graphic art in Figs 1-2.

## Author contributions

CER directed the project, originated the hypothesis, and wrote the first draft of the manuscript. DZ led the statistical analysis and contributed to manuscript preparation and revision. MSH managed the data and images, performed annotations and image analysis, and contributed to manuscript preparation. AS performed the MP-IHC experiments, and JJ performed MP-IHC post-acquisition image processing and analysis. GSK and XL contributed to antibody and target optimization and revised the manuscript. HCM, MAC, and RMF led the collection of locus coeruleus specimens and related pathological assessment and contributed to manuscript revision. DM led the MP-IHC experiments and contributed to hypothesis refinement and manuscript preparation and revision.

## Data availability

The datasets and code are available from the corresponding author upon reasonable request.

## Notes

### Author Declarations

All BBDP, UA-HBB, and UK-ADRC subjects provided written consent for study procedures, autopsy and sharing of de-identified data prior to enrollment. The study and its consenting procedures were approved by the Western IRB of Puyallup, Washington [BBDP], University of Auckland Human Participants Ethics Committee [UA-HBB], and University of Kentucky IRB [UK-ADRC] gave ethical approval of this work.

