## supplement for "ApoER2-Dab1 disruption as the origin of pTau-related neurodegeneration in sporadic Alzheimer’s disease"

### I. Supplementary Tables

**Table S1. Competing explanatory hypotheses: Tau prion-like connectome-based spread vs dendritic RAAAD-P-LTP pathway disruption**

**Table S2. Sixty-four cases spanning the clinicopathological spectrum of sAD**

**Table S3. Key resources**

**Table S4. False discovery rate adjusted p-values for manuscript figures**

**Table S1. Competing explanatory hypotheses: Tau prion-like connectome-based spread vs dendritic RAAAD-P-LTP pathway disruption**

| | Tau prion-like connectome-based spread hypothesis | Dendritic RAAAD-P-LTP disruption hypothesis |
|---|---|---|
| **Overview & Predictions** | | |
| | • Pathogenic Tau is the underlying cause of neurodegeneration<br>• Tau has unique prion-like features—including the ability to serve as bio-template to convert native Tau into pathogenic species—that enable neuron-to-neuron spread<br>• Progression reflects connectome-based spread throughout the brain from a single point of origin<br>• Tau accumulation is the primary histopathological feature | • Dendritic ApoER2-Dab1 disruption is the underlying cause of neurodegeneration<br>• ApoER2-Dab1 disruption traps ApoER2 ligands in the extracellular space and disrupts signaling pathways that mediate cytoskeletal and synaptic stability<br>• pTau is locally generated by ApoER2 expressing neurons in response to ApoER2-Dab1 disruption<br>• High ApoER2 expression and demand for ApoER2-Dab1 activation predispose specific neurons to pTau accumulation<br>• pTau is only one of multiple RAAAD-P-LTP pathway components that accumulate together in response to ApoER2-Dab1 disruption |
| **Global considerations** | | |
| **Mechanistic links to cytoskeletal and synaptic dysfunction and memory deficits** | | |
| Formation of new memories requires fine-tuned control of molecular pathways that shape and strengthen the actin cytoskeleton, microtubule cytoskeleton, and receptor complexes located within the dendrites of excitatory neurons (**Fig 2B**) | No clear mechanism to explain destabilization of the actin cytoskeleton or postsynaptic receptor complexes | Provides a straightforward mechanism—dendritic ApoER2-Dab1 disruption—that can explain destabilization of the actin and microtubule cytoskeletons, synaptic dysfunction, and cognitive deficits that characterize sAD in humans (**Fig 2C**) |
| **Intrinsic molecular features to explain local production of pTau** | | |
| Experimental evidence indicates that disruption of the ApoER2-Dab1 pathway induces Tau hyperphosphorylation [1-5] | Does not propose a mechanism for local production | **Finding that ApoER2 expression parallels laminar and cellular distribution of pTau aligns with RAAAD-P-LTP disruption hypothesis**<br>ApoER2 is strongly expressed by each of five sampled neuron populations that are vulnerable to NFT pathology (**Fig 3**) |
| **Connectome-based pTau spread vs. local production at multiple anatomical locations** | | |
| Rodent and cellular models indicate that trans-synaptic Tau transmission is possible [6] | Discrepancies between neuronal connectivity and NFT progression are extensive and detailed in each section below<br>Accommodation requires hypothetical projections and a revised model of brain connectivity [7-9] | **Posits that pTau is locally produced by ApoER2-expressing neurons, thus no assumptions about neuronal connectivity are needed**<br>Does not require hypothetical projections or a revised model of brain connectivity |
| **Co-accumulation of RAAAD-P-LTP components in affected regions** | | |
| pTau is one of multiple RAAAD-P-LTP components that accumulate together in affected regions (**Figs 4-9**), including:<br>• Extracellular ApoER2 ligands (ApoE, ApoJ)<br>• Neuronal ApoER2 signaling partners:<br>○ upstream to pTau (Dab1, pP85α)<br>○ parallel to pTau (pLIMK1, pPSD95) | No clear mechanism to explain why multiple RAAAD-P-LTP components—both upstream and parallel to pTau production—accumulate together with pTau<br>-Since prion-like properties that enable propagation are thought to be unique to Tau, connectome-based spread of multiple RAAAD-P-LTP components is not likely | **Finding that pTau is one of numerous RAAAD-P-LTP components that accumulate together with pTau aligns with RAAAD-P-LTP disruption hypothesis**<br>• Demonstrates expression of molecular machinery required for local pTau production<br>• Accumulation of RAAAD-P-LTP components that are both upstream and parallel to pTau production aligns with concept of ApoER2-Dab1 disruption |
| **Dendritic origin of pTau lesions** | | |
| pTau pathology originates in distal dendritic tips with sequential progression to proximal dendrites, soma, and finally axons [7 9 10]<br>ApoER2, Dab1 and downstream RAAAD-P-LTP signaling partners are localized to PI-enriched lipid rafts within dendritic spines [11-15] | Origin of pTau lesions in distal dendrites is attributed to axon-to-dendrite spread<br>No mechanism to explain why multiple RAAAD-P-LTP components accumulate together with pTau in dystrophic dendrites | **Finding that RAAAD-P-LTP components including pTau accumulate in dendrites aligns with dendritic RAAAD-P-LTP disruption hypothesis**<br>ApoER2 is strongly expressed within dendritic projections emanating from vulnerable neuron populations (**Fig 3**)<br>Multiple RAAAD-P-LTP components co-accumulate in MAP2-labeled dystrophic dendrites in each region (**Figs 4-9**) |
| **Mechanistic & spatial links between four pTau-containing lesions** | | |
| Molecular mechanisms linking four pTau-containing sAD lesions (NTs, NFTs, NPs, GVDs) are not yet clearly defined | No clear mechanism to spatially link NT/NFTs to NPs and GVDs | Provides a straightforward mechanism—dendritic ApoER2-Dab1 disruption—that could mechanistically and spatially link these four pTau-containing lesions (**Fig 2C**) |

| | Tau prion-like connectome-based spread hypothesis | Dendritic RAAAD-P-LTP disruption hypothesis |
|---|---|---|
| **Region-specific considerations** | | |
| **Entorhinal cortex (ErC)** | | |
| **Laminar, cellular, and subcellular pTau distribution** | | |
| • ErC L2 stellate & pyramidal neurons are the first cortical neurons to accumulate pTau (NFT I)<br>• Followed by L4 pyramids (NFT II)<br>• L3 neurons are spared until later stages<br>• pTau pathology originates in distal dendrites, followed by cell bodies and finally axons [7,9,16] | **No clear explanation for this laminar and cellular distribution of pTau** | **Finding that ApoER2 expression parallels laminar and cellular distribution of pTau aligns with RAAAD-P-LTP disruption hypothesis**<br>• ApoER2 is strongly expressed by ErC L2 stellate neurons and subsets of L2 and L4 pyramids (**Fig 3**)<br>• ApoER2 is strongly expressed within basal & apical dendritic tufts emanating from L2 neurons (**Fig 3**)<br>• The strong expression in L2 and low expression in L3 creates a visible laminar threshold at the L2-L3 border (**Fig 3**) |
| **pTau spread: neuronal connectivity** | | |
| • pTau lesions are classically thought to originate in the LC before progressing specifically to ErC L2.[17,18]<br>• LC axons arborize over large areas and functionally diverse targets,[19-24] including pyramidal neurons in all layers of the human temporal cortex (**Fig 1**). [25]<br>• ErC pTau pathology precedes LC in some cases [26] | **Discrepancies with neuronal connectivity**:<br>-No known connections or mechanisms to explain selective spread of pTau from LC to ErC L2 while sparing other LC-connected layers and neurons | **No assumptions about neuronal connectivity are needed** |
| **ProS-CA1 border region** | | |
| **Laminar, cellular, and subcellular pTau distribution** | | |
| -ProS-CA1 border region is the first hippocampal area to accumulate pTau (NFT stage II)<br>-Involvement of the basal 'stripe' of the ProS is particularly prominent and early<br>-pTau later accumulates the apical stripe | **No clear explanation for this laminar and cellular distribution of pTau** | **Finding that ApoER2 expression parallels laminar and cellular distribution of pTau aligns with RAAAD-P-LTP disruption hypothesis**<br>- ApoER2 is strongly expressed by two basal ProS-CA1 neuron populations (basal pyramids and CR-like cells that reside in the basal stripe) (**Fig 3**)<br>- Moderate-to-strong expression in basal & apical dendritic tufts emanating from ProS-CA1 pyramids (**Fig 3**) |
| **pTau spread: neuronal connectivity** | | |
| -In NFT stage I, pTau pathology is confined to ErC L2 projection neurons. In NFT stage II, pTau pathology progresses to include the basal lamina of the ProS-CA1 border region. In successive NFT stages, pTau pathology progresses throughout the cornu ammonis. DG neurons are spared from NFT pathology until late-stage sAD (NFT stages V-VI) (**Fig 1A**).<br>-The first connection of the unidirectional tri-synaptic memory circuit is made between axons projecting from ErC L2 neurons to dendritic arbors emanating from DG neurons (**Fig 1A**).[27] These axons are known as the 'perforant path' because they perforate (bypass) the subiculum before synapsing with DG neurons.[27] The second and third synapses underlying memory connect DG neurons to CA3 pyramids (via mossy fiber axons) and CA3 neurons to CA1 pyramids (via Schaffer collateral axons), respectively.<br>- Thus, NFT progression proceeds in a direction opposite to unidirectional connectivity in the medial temporal lobe memory system [27,28] and spares the major synaptic recipient of ErC L2 neurons (DG neurons) until late-stage sAD (**Fig 1A**).[7] | **Discrepancies with neuronal connectivity**:<br>-Since cortical NFT pathology begins in ErC L2, a connectome-based spread model predicts sequential pTau spread in accordance with unidirectional connectivity in the medial temporal lobe memory system (from ErC L2 to DG, followed by DG to CA3, CA3 to CA1, and finally to the subiculum).<br><br>**There are no known connections or mechanisms to explain**:<br>-Why NFT progression proceeds in a direction *opposite* to unidirectional connectivity in the medial temporal lobe memory system<br>-Selective spread of pTau from ErC L2 (NFT stage I) to ProS (NFT stage II) [27,28] while sparing the major synaptic recipient of ErC L2 neurons (DG neurons) until end-stage sAD (Fig 1A).[7] | **No assumptions about neuronal connectivity are needed** |

| | Tau prion-like connectome-based spread hypothesis | Dendritic RAAAD-P-LTP disruption hypothesis |
|---|---|---|
| **Solitary L5 & L2/3 temporal pyramids** | | |
| **Cellular and laminar pTau distribution** | | |
| -pTau accumulates in distal dendrites of rare, isolated L5 and L2/3 neocortical pyramids in the earliest stage of sAD (NFT stage 1)<br>-all dendrites of each affected neuron are generally involved while neighboring neurons are spared<br>-Spiny stellate cells in L4 are spared even in advanced sAD[9 29] | **No clear explanation for this laminar and cellular distribution of pTau** | **Finding that ApoER2 expression parallels cellular and laminar distribution of pTau in neocortex aligns with RAAAD-P-LTP disruption hypothesis**<br>- ApoER2 strongly expressed by subset of L5 and L2/3 pyramidal neurons, their basal and apical dendrites, and dendritic tufts in the L2/3 border region (**Fig 3**)<br>-L4 spiny stellate cells have weak or absent ApoER2 expression (**Fig 3**)<br>-The strong expression in L2 and low expression in L3 creates a visible laminar threshold at the L2-L3 border (**Fig 3)** |
| **pTau spread: neuronal connectivity** | | |
| -pTau lesions are classically thought to originate in the LC before spreading to the cortex.[17 18]<br>-LC axons arborize over large areas and functionally diverse targets,[19-24] including pyramidal neurons in all layers of the human temporal cortex (**Fig 1**). [25]<br>-Individual projection neurons classically innervate hundreds (or even thousands) of neighboring target neurons [30 31] rather than making one-to-one connections with a single target neuron. Yet pTau accumulates within solitary L5 and L2/3 neocortical pyramids in very early sAD stages (**Fig 1A**)[9 32], while sparing neighboring neurons<br>- pTau accumulations are evident in numerous dendritic tips emanating from the same, solitary pyramidal neurons | **Discrepancies with neuronal connectivity**:<br>-No known connections or mechanisms to explain:<br>-selective pTau spread to L5 & L2/3 while sparing other layers.<br>-selective spread of pTau to isolated pyramids while sparing neighboring neurons<br>The observation that all dendrites of affected solitary pyramids accumulate pTau while neighboring neurons are spared is not easily reconciled with prion-like propagation<br>Braak et al [8 9] suggested we may need to rethink traditional brain connectivity in order to reconcile these isolated pTau lesions with the Tau prion-like spread hypothesis. In this connectome-inspired revision, all pre-synaptic terminals emanating from a single donor axon synapse on dendrites emanating from a single target neuron and refrain from contacting neighboring neurons.[8 9] | **No assumptions about neuronal connectivity are needed**<br>Finding that pTau accumulates within multiple distal dendrites of rare, isolated L5 and L2/3 neocortical pyramids is attributed to ApoER2-Dab1 disruption—with ensuring Tau hyperphosphorylation—in ApoER2-enriched dendritic spines of L5 and L2/3 pyramids |
| **Pontine LC-PC and Raphe nucleus** | | |
| **pTau distribution** | | |
| -Pre-tangles are evident in the LC and peri-coeruleus in the earliest stages of sAD (pre-tangle stages a/b)<br>-LC fusiform-shaped neurons prominently affected<br>-pTau later accumulates in raphe nucleus (pre-tangle stage c)<br>-Classically precedes cortical pTau; however, ErC precedes LC and raphe nucleus in some cases [26] | **No clear explanation for this laminar and cellular distribution of pTau** | **Finding that ApoER2 expression matches the cellular distribution of pTau aligns with RAAAD-P-LTP disruption hypothesis**<br>- ApoER2 is strongly expressed by LC and raphe nucleus neurons including fusiform shaped projection neurons, and neurons located between these two nuclei (**Fig 3**)<br>-ApoER2 is strongly expressed in highly branched neuritic projections emanating from LC and raphe neurons and in the peri-coeruleus region harboring MAP2-labeled dendritic tufts emanating from LC neurons (**Fig 3**)<br>-ApoER2 is also strongly expressed within proximal dendrites, and dense dendritic projections emanating into in the peri-coeruleus (**Fig 3**) |
| **pTau spread: neuronal connectivity** | | |
| -LC axons arborize over large areas and functionally diverse targets,[19-24] including pyramidal neurons in all layers of the human temporal cortex (**Fig 1**) [25]<br>-Raphe nucleus serotonergic axons arborize over large areas and functionally diverse targets [33] | **Discrepancies with neuronal connectivity**:<br>-No known connections or mechanisms to explain selective spread from LC and/or the raphe nucleus to either ErC L2 neurons or rare, isolated L5 and L2/3 neocortical pyramids | **No assumptions about neuronal connectivity are needed** |

**Table S2 – Part 1. Sixty-four cases spanning the clinicopathological spectrum of sAD**

| Gender | Age (years) | PMI (hours) | NFT stage (0-6) | Amyloid plaques (0-15) [a] | Neuritic plaques (0-3) | MMSE (0-30) | ApoE status |
|---|---|---|---|---|---|---|---|
| **Cohort 1: Brain and Body Donation Program (BBDP) at the Banner Sun Health Research Institute (n=34)** | | | | | | | |
| **Alzheimer's Disease** | | | | | | | |
| Female | 85-89 | 3.4 | 6 | 14 | 3 | 2 | 3/3 |
| Male | 70-74 | 4.8 | 6 | 14.5 | 3 | 7 | 3/4 |
| Male | 75-79 | 3.6 | 6 | 15 | 3 | 10 | 3/4 |
| Female | 70-74 | 4.9 | 6 | 15 | 3 | 17 | 3/4 |
| Female | 80-84 | 4.0 | 6 | 15 | 3 | 19 | 3/3 |
| Female | 80-84 | 3.3 | 6 | 15 | 3 | 24 | 3/3 |
| Female | 75-79 | 3.1 | 5 | 15 | 3 | 6 | 3/3 |
| Female | 90+ | 2.2 | 5 | 15 | 3 | 13 | 3/3 |
| Male | 80-84 | 4.0 | 5 | 14 | 3 | 14 | 3/4 |
| Male | 85-89 | 2.2 | 5 | 12.5 | 3 | 21 | 2/3 |
| **Mild Cognitive Impairment** | | | | | | | |
| Female | 90+ | 3.2 | 4 | 5 | 3 | 22 | 3/3 |
| Male | 85-89 | 4.2 | 4 | 0 | 0 | 23 | 3/3 |
| Female | 75-79 | 3.2 | 4 | 4.5 | 0 | 24 | 2/3 |
| Female | 90+ | 3.2 | 4 | 13.5 | 3 | 26 | 2/3 |
| Female | 85-89 | 3.6 | 4 | 10 | 3 | 28 | 2/3 |
| Female | 80-84 | 3.0 | 4 | 12.5 | 3 | 29 | 2/3 |
| Female | 80-84 | 3.2 | 4 | 11 | 3 | 29 | 2/3 |
| Male | 85-89 | 2.2 | 3 | 8.25 | 2 | 28 | 3/3 |
| Male | 80-84 | 2.0 | 1 | 0.25 | 0 | | 3/3 |
| **Age-matched Controls** | | | | | | | |
| Female | 90+ | 3.0 | 3 | 6.5 | 1 | 27 | 3/3 |
| Male | 70-74 | 3.5 | 3 | 0 | 0 | 27 | 3/3 |
| Female | 85-89 | 3.1 | 3 | 0 | 0 | 28 | 3/4 |
| Female | 75-79 | 2.5 | 2 | 0 | 0 | 28 | 3/3 |
| Male | 90+ | 3.4 | 1 | 0.5 | 1 | 27 | 3/3 |
| Male | 75-79 | 2.3 | 1 | 5.5 | 1 | 29 | 3/4 |
| Female | 80-84 | 2.1 | 1 | 1 | 1 | 29 | 2/3 |
| Male | 70-74 | 4.6 | 1 | 0 | 0 | 29 | 3/3 |
| Male | 90+ | 3.0 | 1 | 0 | 0 | 30 | 3/3 |
| Male | 85-89 | 3.0 | 1 | 4.25 | 1 | | 3/3 |
| **Middle-age Controls** | | | | | | | |
| Male | 60-64 | 2.3 | 1 | 0.5 | 1 | | 3/3 |
| Female | 55-59 | 3.1 | 1 | 1 | 0 | | 3/3 |
| Female | 50-54 | 4.7 | 1 | 0 | 0 | | 3/3 |
| Male | 35-39 | 3.0 | 0 | 0 | 0 | | 3/3 |
| Male | 45-49 | 4.5 | 0 | 0 | 0 | | 3/3 |

#### Table S2 – Part 2. Sixty-four cases spanning the clinicopathological spectrum of sAD

| Gender | Age (years) | PMI (hours) | NFT stage (0-6) | Thal phase (0-5) | Neuritic plaques (0-3) | MMSE (0-30) | ApoE status |
|---|---|---|---|---|---|---|---|
| **Cohort 2: Neurological Foundation Human Brain Bank at the University of Auckland, New Zealand (n=18)** | | | | | | | |
| **Alzheimer's Disease** | | | | | | | |
| Female | 60-64 | 16.0 | 6 | 5 | 2 | | 4/4 |
| Male | 65-69 | 12.0 | 5 | 5 | 2 | | 3/3 |
| Male | 70-74 | 5.0 | 5 | | 3 | | 3/3 |
| Female | 70-74 | 8.5 | 5 | 5 | 2 | | 3/3 |
| Male | 75-79 | 11.5 | 3 | 5 | 3 | | 3/3 |
| Female | 80-84 | | 5 | 4 | 2 | | 3/4 |
| Male | 80-84 | 15.0 | 5 | | 2 | | 3/4 |
| Male | 85-89 | | 4 | 3 | 1 | | 3/4 |
| Male | 85-89 | 24.0 | 4 | 4 | 2 | | 3/4 |
| Female | 90-94 | 11.5 | 6 | 5 | 2 | | 3/4 |
| **Age-matched Controls** | | | | | | | |
| Male | 55-59 | 24.5 | 1 | | | | 3/3 |
| Male | 60-64 | 9.0 | 1 | | | | 3/3 |
| Male | 70-74 | 23.0 | 1 | | | | |
| Male | 70-74 | 5.5 | 2 | | | | 3/3 |
| Male | 70-74 | 13.0 | 2 | | | | 3/3 |
| Male | 75-79 | 13.0 | 2 | | | | 3/3 |
| Male | 75-79 | 23.0 | 1 | | | | 3/3 |
| Female | 75-79 | 20.0 | 3 | | | | 3/3 |
| **Cohort 3: University of Kentucky Alzheimer's Disease Research Center (UK-ADRC) Biobank (n=12)** | | | | | | | |
| **Gender** | **Age (years)** | **PMI (hours)** | **NFT stage (0-6)** | **Thal phase (0-5)** | **Neuritic plaques (0-3)** | **MMSE (0-30)** | **ApoE status** |
| **Alzheimer's Disease** | | | | | | | |
| Male | 75-79 | 3.3 | 6 | | Frequent | 5 | 4/4 |
| Male | 80-84 | 3.3 | 6 | 5 | Frequent | 2 | 3/4 |
| Female | 85-89 | 2.2 | 6 | 5 | Frequent | 10 | 2/3 |
| Female | 90-94 | 2.8 | 6 | | Frequent | 0 | 3/3 |
| **Mild Cognitive Impairment** | | | | | | | |
| Male | 80-84 | 3.5 | 4 | | Moderate | 24 | 3/4 |
| Female | 80-84 | 2.5 | 2 | 0 | None | 26 | 3/3 |
| Male | 85-89 | 2.8 | 4 | | None | 27 | 3/3 |
| Female | 90-94 | 2.3 | 3 | 3 | None | 28 | 3/4 |
| **Age-matched Controls** | | | | | | | |
| Male | 70-74 | 2.6 | 0 | 3 | None | 28 | 3/3 |
| Female | 80-84 | 2.8 | 1 | 1 | None | 30 | 3/4 |
| Male | 85-89 | 4.0 | 2 | 2 | None | 28 | 3/3 |
| Female | 90-94 | 1.3 | 1 | | None | 24 | 3/3 |

#### Table S3. Key resources

| *Reagent type or resource* | *Target* | *Designation* | *Type* | *Source or reference* | *Catalog #* | *Previous Use* | *Comment* |
|---|---|---|---|---|---|---|---|
| **RNA probes** | | | | | | | |
| ISH probe | ApoER2 mRNA probe 1 | LRP8a | ISH probe | ACD, Biotechne | 807461 | ISH human brain | |
| ISH probe | ApoER2 mRNA probe 2 | LRP8b | ISH probe | ACD, Biotechne | 1160541 | n/a | |
| ISH probe | Reelin mRNA probe | RELN | ISH probe | ACD, Biotechne | 413051 | n/a | |
| **Primary antibodies** | | | | | | | |
| antibody | ApoER2 | ApoER2 | Rabbit IgG | Millipore-Sigma | SAB2103110 | IHC human brain, WB | IHC 1:80-1:150 |
| antibody | Disabled homolog 1 | Dab1 | Rabbit IgG | Invitrogen | PA5-86617 | IHC human brain, WB | IHC 1:50 |
| antibody | Tyr607-phosphorylated p85α | $pP85\alpha_{Tyr607}$ | Rabbit IgG | Invitrogen | PA5-104853 | IHC human brain, WB | IHC 1:100-1:150 |
| antibody | Thr508-phosphorylated LIM kinase-1 | $pLIMK1_{Thr508}$ | Rabbit IgG | Invitrogen | PA5-104925 | IHC human brain, WB | IHC 1:100 |
| antibody | Thr19-phosphorylated PSD95 (DLG4) | $pPSD95_{Thr19}$ | Rabbit IgG | Millipore-Sigma | ABN998 | IHC human brain, WB | IHC 1:50-1:100 |
| antibody | Ser202/Thr205-phosphorylated Tau | pTau | Mouse IgG1 [AT8] | Invitrogen | MN1020 | IHC human brain, WB | IHC 1:100 |
| antibody | ApoE | ApoE | Mouse IgG1 [WUE4] | Novus Biologicals | NB110-60531 | IHC human brain, WB | IHC 1:60-1:100 |
| antibody | ApoE | ApoE | Chicken IgG | New England Peptide | n/a | IHC human brain, WB | IHC 1:50-1:100 |
| antibody | ApoJ | ApoJ | Rabbit IgG | Invitrogen | PA5-24426 | IHC human brain, WB | 1:100-1:150 |
| antibody | Reelin | Reelin | Mouse IgG2a [E-5] | Santa Cruz | SC-25346 | IHC human brain, WB | IHC 1:50 |
| antibody | Amyloid Beta Protein | Aβ | Mouse IgG2b [MOAB-2] | Novus Biologicals | NBP2-13075 | IHC human brain, WB | IHC 1:100 |
| antibody | Tyr220-phosphorylated Dab1 | $Dab1_{Tyr220}$ | Rabbit IgG | Invitrogen | PA5-104586 | IHC human, WB | IHC 1:50 |
| antibody | LRP1 | LRP1 | Mouse IgG1 [A2MR-beta1] | Invitrogen | 37-7600 | IHC human, WB | IHC 1:100 |
| antibody | VLDLR | VLDLR | Mouse IgG1 | Diagnocine [VL1A9] | BML031 | IHC human brain, WB | IHC 1:150-1:200 |
| antibody | Neuronal marker NeuN (antibody 1) | NEUN | Guinea Pig IgG | Millipore Sigma | ABN90P | IHC human brain, WB | IHC 1:100 |
| antibody | Microtubule associated protein 2 | MAP2 | Mouse IgG3 [885232] | R&D Systems | MAB8304 | IHC human brain, WB | IHC 1:100 |
| antibody | Neurofilament light chain | NFL | Mouse IgG1 [NFL2] | Biolegend | 846002 | IHC human brain, WB | IHC 1:100 |
| antibody | Neurofilament light chain | NFL | Mouse IgG1 [NFL3] | Biolegend | 845902 | IHC human brain, WB | IHC 1:100 |
| antibody | Synaptophysin | SYNAP | Mouse IgM [SP15] | Millipore Sigma | MAB328 | IHC human brain, WB | IHC 1:100 |
| **Secondary Antibodies** | | | | | | | |
| antibody | Goat anti-Rat IgG | | | Jackson | 112-035-167 | | 1.6 ug/mL |
| antibody | Goat anti-Mouse IgG1 | | | Jackson | 115-035-205 | | 1.6 ug/mL |
| antibody | Goat anti-Mouse IgG2a | | | Jackson | 115-035-206 | | 1.6 ug/mL |
| antibody | Goat anti-Mouse IgG2b | | | Jackson | 115-035-207 | | 1.6 ug/mL |
| antibody | Goat anti-Mouse IgG3 | | | Jackson | 115-035-209 | | 1.6 ug/mL |
| antibody | Goat anti-Mouse IgM | | | Jackson | 115-035-075 | | 1.6 ug/mL |
| antibody | Donkey anti-Rabbit IgG | | | Jackson | 711-035-152 | | 1.6 ug/mL |
| antibody | Donkey anti-Chicken IgY | | | Jackson | 703-035-155 | | 1.6 ug/mL |

Abbreviations: ISH, In situ hybridization; IHC, Immunohistochemistry; WB, western blot

#### Table S4. False discovery rate adjusted p-values for manuscript figures [a]

| | by Group [b] | | vs NFT Stage [c] | | vs MMSE [c] | |
|---|---|---|---|---|---|---|
| | **p-value** | **Sharpened q-value** | **p-value** | **Sharpened q-value** | **p-value** | **Sharpened q-value** |
| **Figure 3.A. ErC** | | | | | | |
| ApoER2 | 0.002 | 0.002 | | | | |
| **Figure 3.C. Temporal Cortex** | | | | | | |
| ApoER2 | 0.002 | 0.002 | | | | |
| **Figure 4.B. ErC** | | | | | | |
| *Row 1* | | | | | | |
| pTau | <0.001 | 0.001 | <0.001 | 0.001 | <0.001 | 0.001 |
| Dab1 | 0.026 | 0.008 | 0.025 | 0.008 | 0.062 | 0.013 |
| pP85α | <0.001 | 0.001 | <0.001 | 0.001 | <0.001 | 0.001 |
| *Row 2* | | | | | | |
| pLIMK1 | 0.006 | 0.004 | 0.002 | 0.002 | <0.001 | 0.001 |
| pPSD95 | <0.001 | 0.001 | <0.001 | 0.001 | <0.001 | 0.001 |
| ApoJ | <0.001 | 0.001 | <0.001 | 0.001 | <0.001 | 0.001 |
| **Figure 6.C. ProS-CA1** | | | | | | |
| *Row 1* | | | | | | |
| Dab1 | 0.001 | 0.002 | <0.001 | 0.001 | <0.001 | 0.001 |
| pTau | <0.001 | 0.001 | <0.001 | 0.001 | <0.001 | 0.001 |
| pPSD95 | <0.001 | 0.001 | <0.001 | 0.001 | <0.001 | 0.001 |
| *Row 2* | | | | | | |
| pP85α | <0.001 | 0.001 | <0.001 | 0.001 | <0.001 | 0.001 |
| pLIMK1 | 0.212 | 0.030 | 0.009 | 0.004 | 0.043 | 0.011 |
| pDab1 | 0.013 | 0.006 | 0.003 | 0.002 | 0.006 | 0.004 |
| **Figure 8.D. Temporal Cortex** | | | | | | |
| Dab1 | 0.005 | 0.003 | <0.001 | 0.001 | 0.043 | 0.011 |
| | | | **vs Amyloid Plaques [c]** | | **vs PSD95 [c]** | |
| | | | 0.018 | 0.007 | 0.001 | 0.002 |
| **Figure 9.C. LC and Raphe Nucleus** | | | | | | |
| | | | **vs NFT Stage [c]** | | | |
| Dab1 | 0.004 | 0.003 | 0.053 | 0.012 | | |
| pTau | <0.001 | 0.001 | <0.001 | 0.001 | | |
| pPSD95 | 0.006 | 0.004 | 0.010 | 0.005 | | |

[a] Based on the two-stage linear step-up procedure described by Benjamini, et al. [1 2].
[b] For Figure 3 & 9, p-values were determined using Wilcoxon rank-sum test by layers. For the other figures, p-values were determined with the Kruskal-Wallis equality-of-populations rank test.
[c] P-values were determined using Spearman's rank correlations test.

### II. Extended Figures

**Ext Fig 3.1. Laminar and cellular patterns of ApoER2 expression in ErC and neocortex**
**Ext Fig 3.2. VLDLR and LRP1 lack the restricted expression observed for ApoER2**
**Ext Fig 4.1 Accumulation of ApoE and ApoJ in the ErC in sAD**
**Ext Fig 5.1 Cytoarchitectural context for early Dab1 accumulation in the ErC**
**Ext Fig 6.1. Multiple RAAAD-P-LTP components accumulate in the ProS-CA1 border region**
**Ext Fig 6.2 Reelin accumulation in the hippocampus and the ProS-CA1 region in sAD**
**Ext Fig 9.1 Dab1 accumulation within plaque-associated dystrophic axons in temporal neocortex in sAD**
**Ext Fig 9.2 Pathological & cytoarchitectural context for neocortical Dab1 accumulation in NFT stage 0**
**Ext Fig 10.1. Intra-neuronal Dab1 inclusions in locus coeruleus and raphe nucleus in sAD**
**Ext Fig 10.2. Extracellular accumulations of ApoER2 ligands in locus coeruleus & raphe nucleus in sAD**

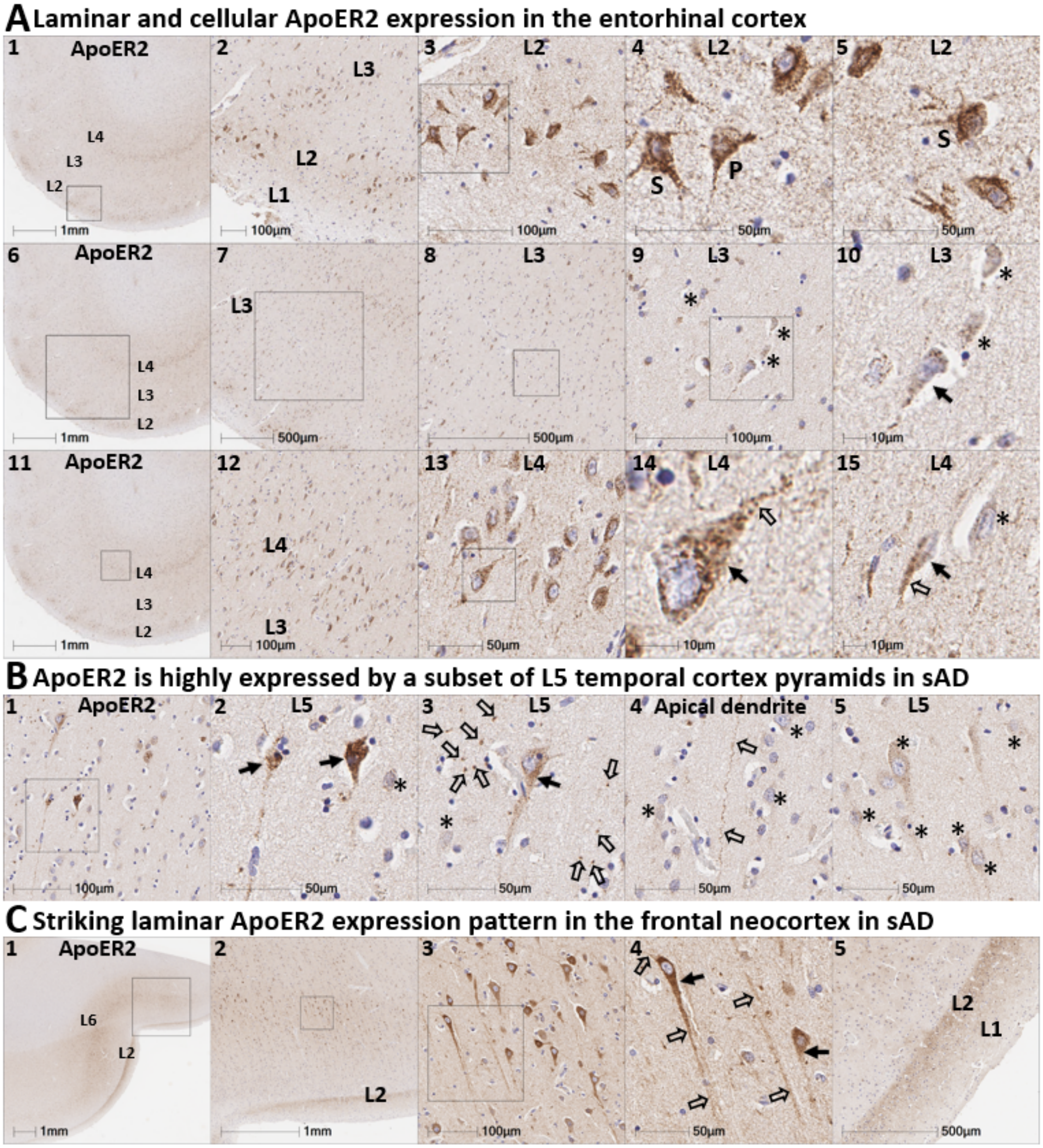


**Ext Fig 3.1. Laminar and cellular patterns of ApoER2 expression in the ErC and neocortex.** Panels **A-C** are coronal sections of the ErC (**A**), middle temporal gyrus (**B**), and frontal cortex (**C**) from one representative middle-aged non-AD control and two sAD cases, respectively. (**A**) ApoER2 is strongly expressed by stellate (demarcated by S in **A**$_{4-5}$) and pyramidal neurons (demarcated by P in **A**$_{4}$) in ErC L2. ApoER2 expression is lower or absent in L3 pyramids (**A**$_{7-10}$) and strongly expressed by a subset of L4 pyramids and surrounding neurites (**A**$_{11-15}$). Examples of L3 and L4 neurons with low ApoER2 expression are demarcated by an * in **A**$_{9,\ 10\ \&\ 15}$. (**B**) ApoER2 is strongly expressed by a subset of neocortical L5 pyramids (solid arrows in **B**$_{2-3}$) and their apical and basal dendritic projections (open arrows in B$_{2-4}$); L5 neurons with low or absent ApoER2 expression are demarcated by an * in **A**$_{4-5}$. (**C**) In the frontal neocortex, ApoER2 is strongly expressed by a subset of L3 and L5 pyramidal neurons and their apical dendritic projections (open arrows in B$_{4}$) and highly-ramified apical dendritic tufts (**C**$_{2,\ 5}$) located in the vicinity of L2.

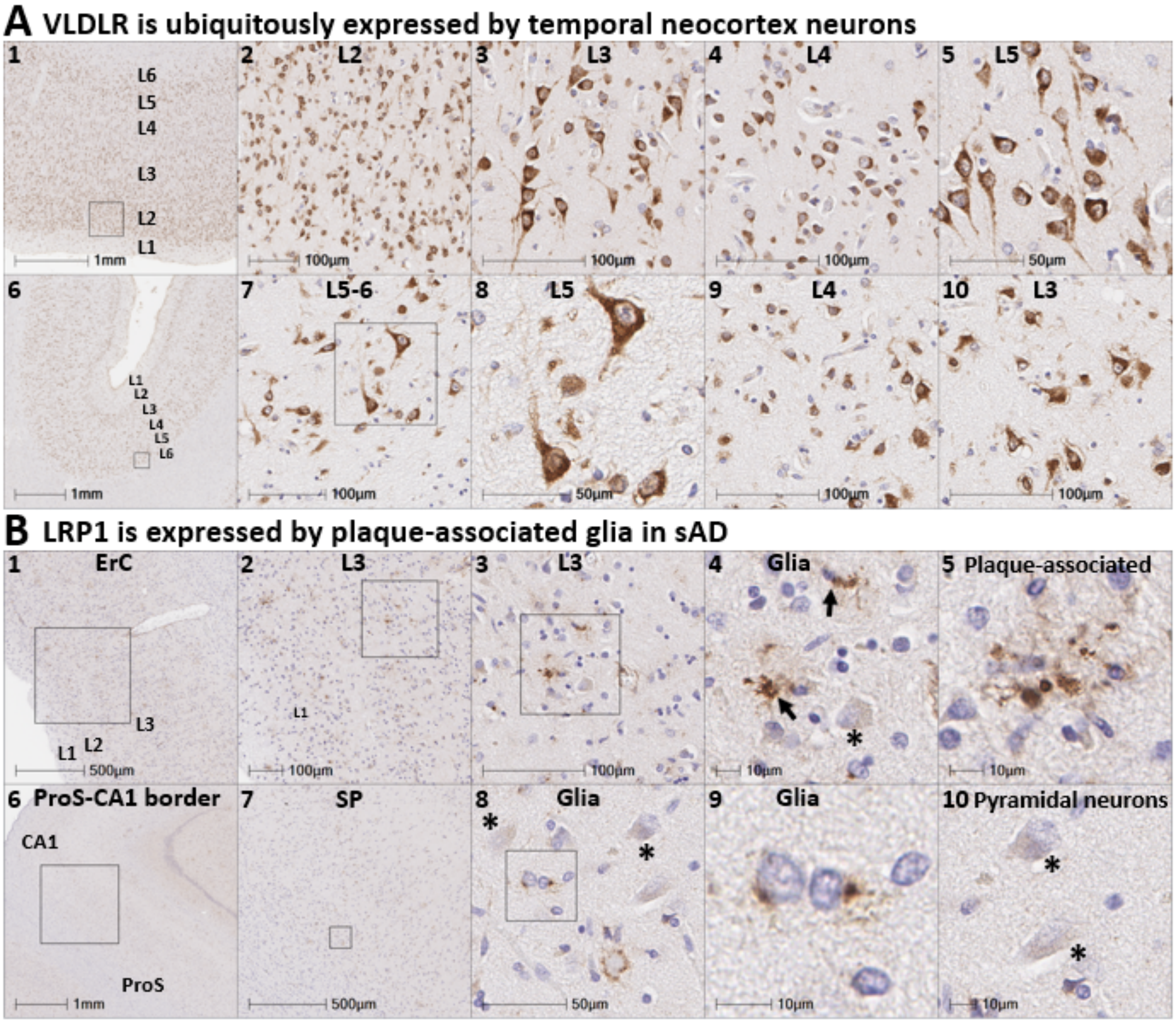


**Ext Fig 3.2. VLDLR and LRP1 lack the restricted expression observed for ApoER2**

Panels **A-B** are coronal sections of the temporal neocortex in a non-AD control (**A$_{1-5}$**) and sAD case (**A$_{6-10}$**), and the ErC (**B$_{1-5}$**) and ProS-CA1 region from a sAD case (**B$_{6-10}$**). The expression of VLDLR and LRP1 was less restricted than ApoER2 and neither closely matched the laminar and cellular distribution of NFT pathology. VLDLR was strongly and ubiquitously expressed by neurons in all neocortical layers including neocortical L4 stellate neurons (**A$_{1-10}$**). LRP1 was expressed by glia and some neurons, with prominent signals in glia surrounding NPs (**B$_{4,\ 8-10}$**). Neurons with low or no LRP1 expression are demarcated by an * in **B$_{4,\ 8\ \&\ 10}$**.

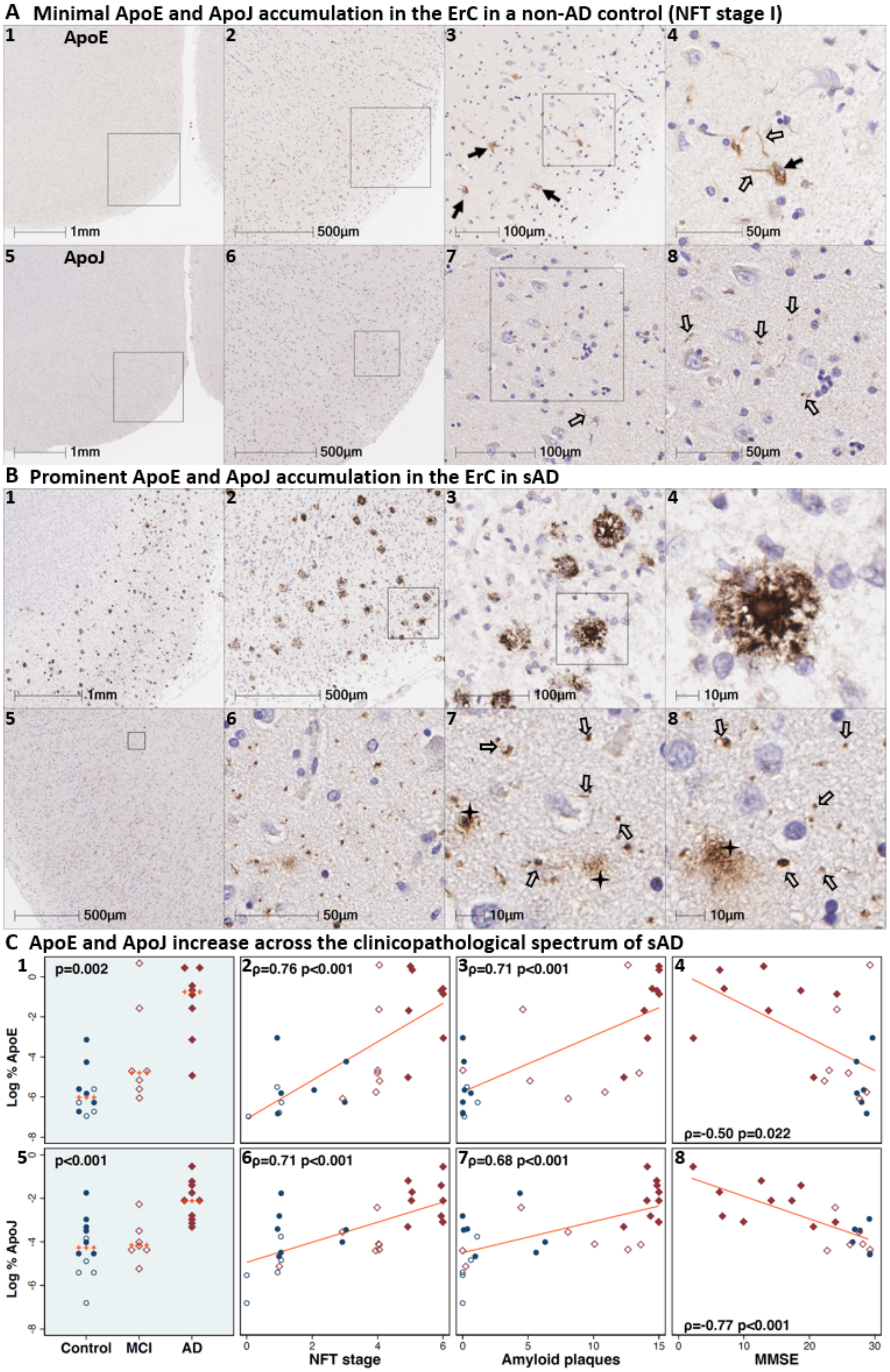


**Ext Fig 4.1 Accumulation of ApoE and ApoJ in the ErC in sAD**

(**A**) Serial coronal sections of the ErC from a non-AD control case in the earliest stage of NFT pathology (NFT stage I) had only subtle ApoE (**A**$_{1-4}$) and ApoJ (**A**$_{5-8}$) expression. (**B**) Prominent extracellular accumulations of ApoE (**B**$_{1-4}$) and ApoJ (**B**$_{5-8}$) were observed in sAD cases. ApoE expression was most prominent in extracellular plaques. ApoJ was evident in both plaques (black stars) and discrete punctae within the neuropil (open arrows in **B**$_{6-8}$). (**C**) ApoE and ApoJ were higher in sAD cases than controls, and positively correlated with histological progression and antemortem cognitive deficits.

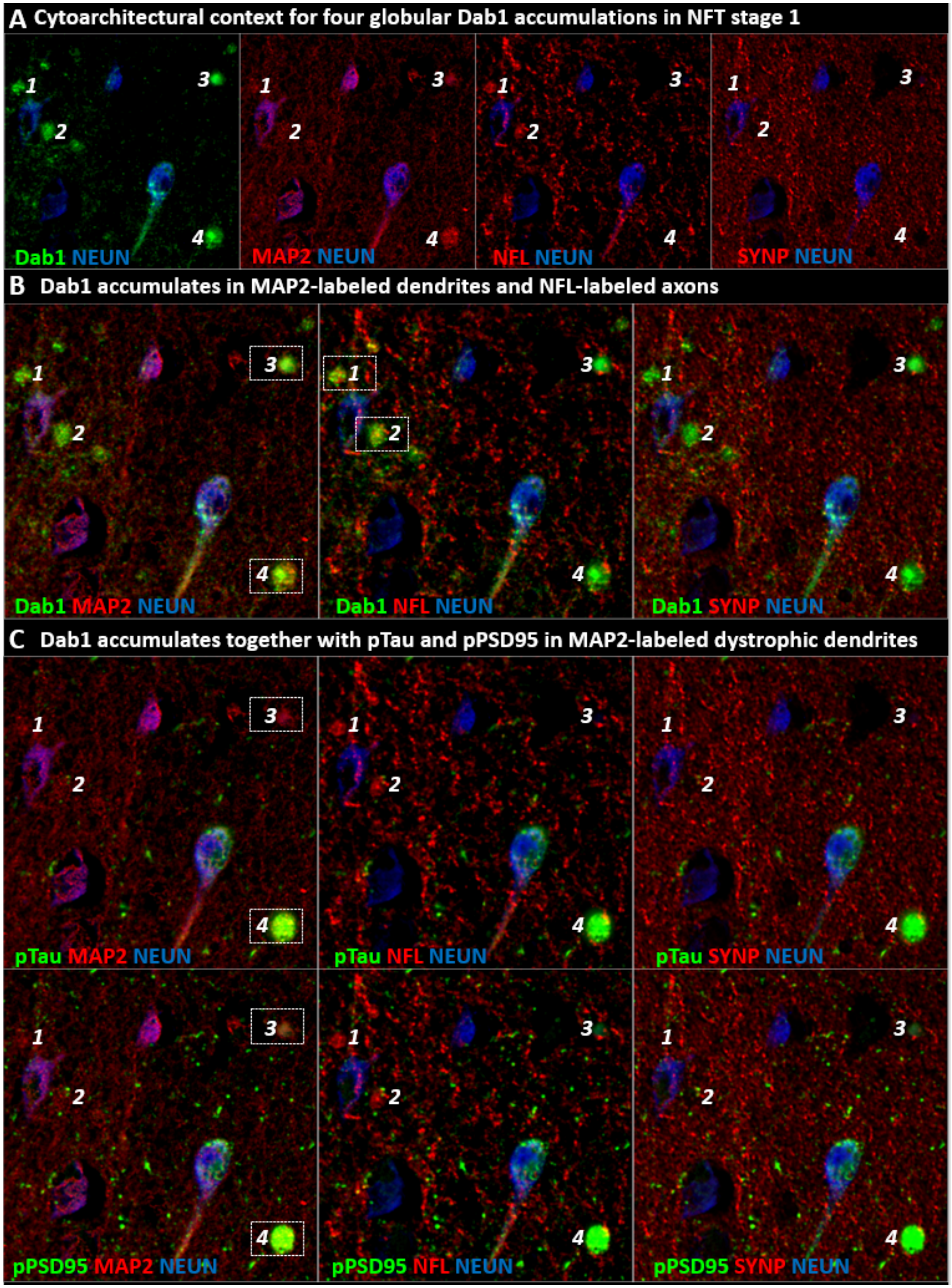


**Ext Fig 5.1 Cytoarchitectural context for early Dab1 accumulation in the ErC**

This figure provides cytoarchitectural context for Dab1 accumulation observed in the NFT stage I case shown in Fig 5. Panel **A** shows Dab1 accumulation in four prominent globular structures (**A$_{1\text{-}4}$**) in the vicinity of affected ErC L2 stellate-shaped and pyramidal neurons. Panel **B** shows that two of these globular Dab1 accumulations colocalized with MAP2-labeled dystrophic dendrites and the other two colocalized with NFL-labeled dystrophic axons. Although these Dab1 structures accumulated in close proximity to synaptophysin (SYNP), no clear co-localization was evident. Panel **C** shows that pTau and pPSD95 expression overlapped with at least one the dendritic Dab1 accumulations but did not appear to colocalize with axonal Dab1.

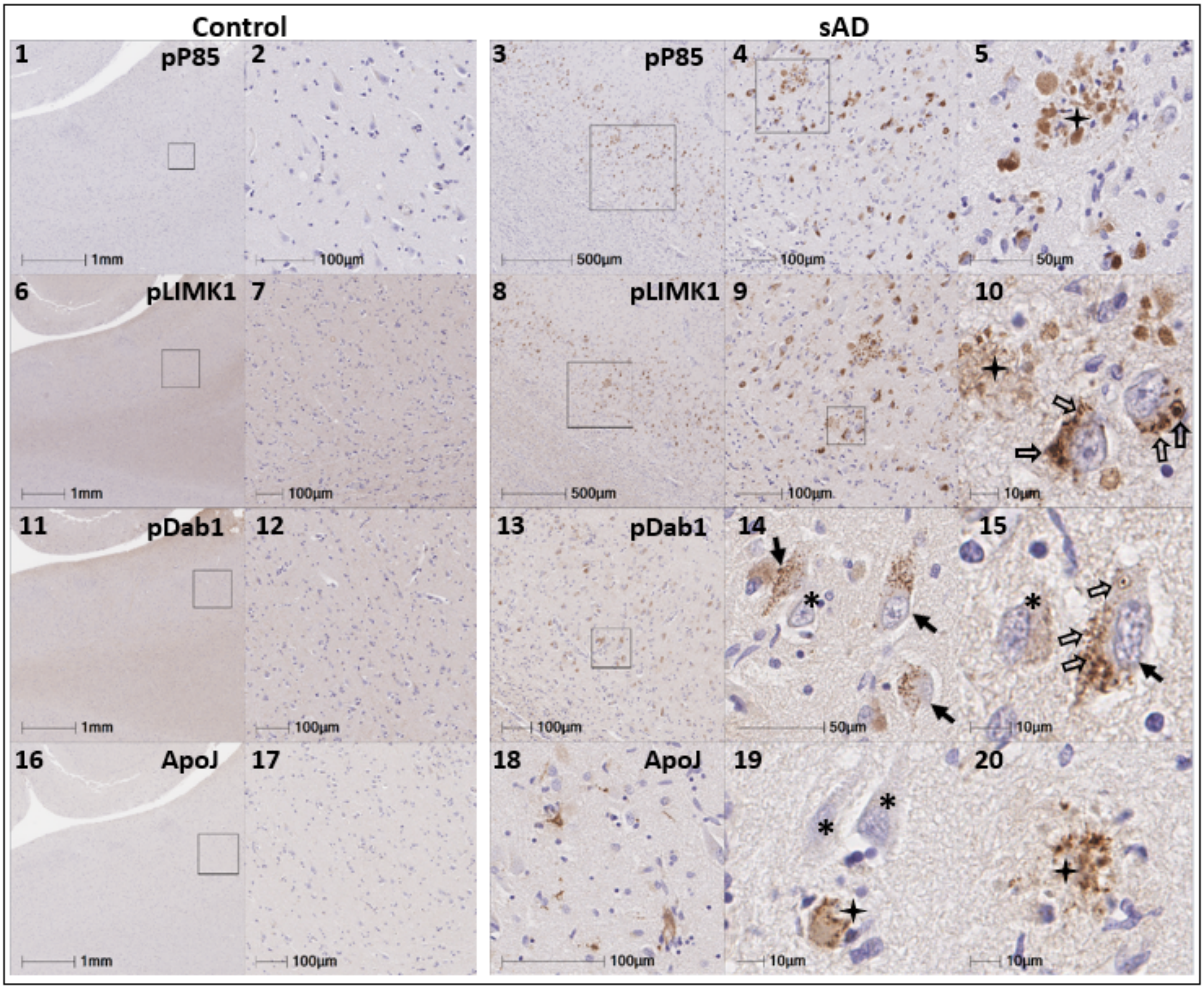


**Ext Fig 6.1. Multiple RAAAD-P-LTP components accumulate in the ProS-CA1 border region**

(**A**) Serial coronal sections of the ProS-CA1 region from representative non-AD control and sAD cases were probed with antibodies targeting RAAAD-P-LTP pathway components (see **Suppl Table 3**). In the non-sAD control (left column), IHC revealed very low expression of pP85α$_{Tyr607}$, pLIMK1$_{Thr508}$, and pDab1$_{Tyr220}$, and ApoJ. By contrast, in sAD (right column), prominent accumulations of pP85α$_{Tyr607}$, pLIMK1$_{Thr508}$, and pDab1$_{Tyr220}$ were observed within abnormal neurons (solid arrows) and in the vicinity of NPs (black stars). Open arrows in **A$_{10}$** and **A$_{15}$** designate granulovacuolar accumulations of pLIMK1$_{Thr508}$ and pDab1$_{Tyr220}$, respectively. ApoJ accumulated primarily in extracellular plaques (black stars). Neighboring neurons with little or no evidence of RAAAD-P-LTP component accumulation are designated with * (**A$_{15, 19}$**).

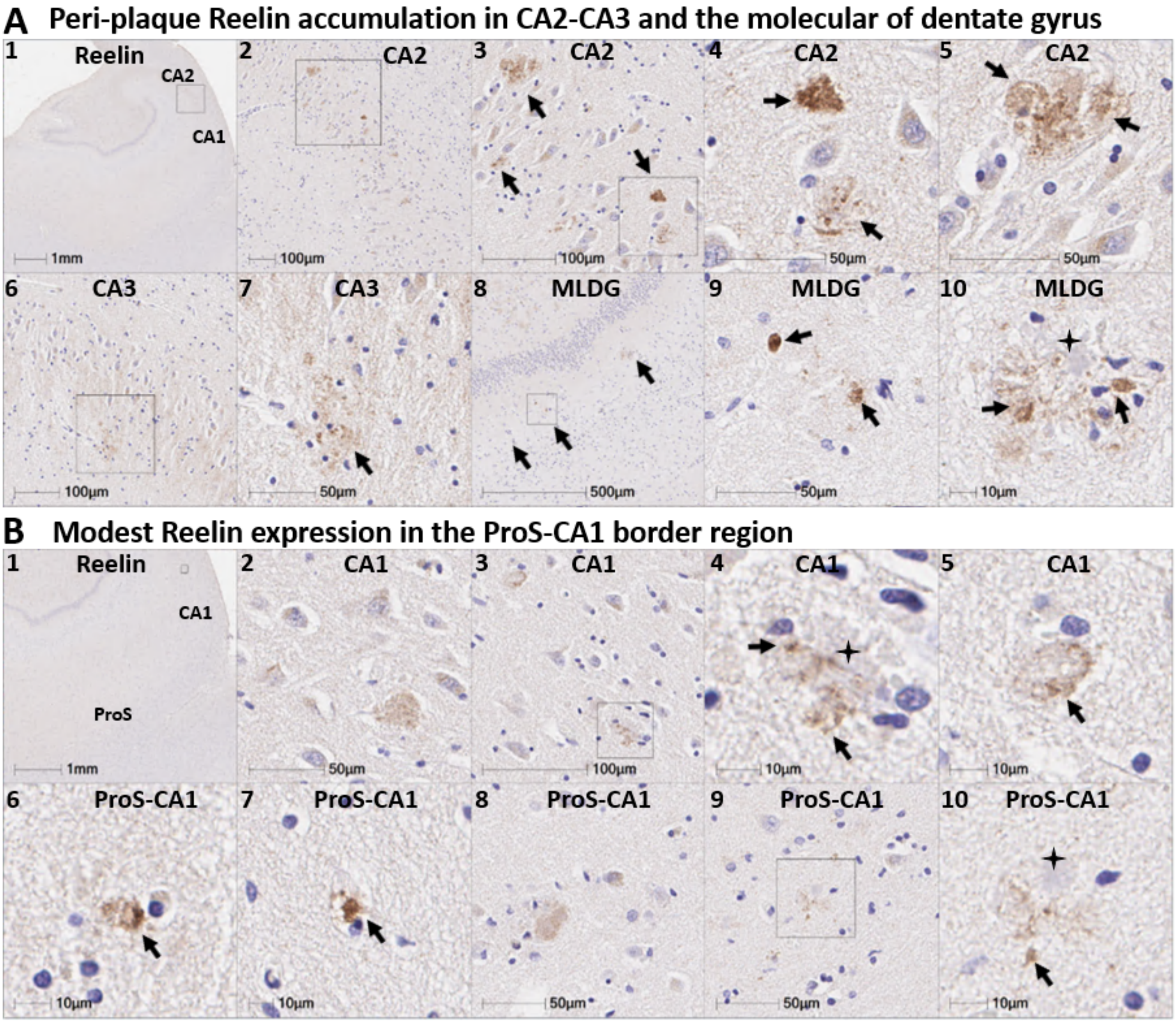


**Ext Fig 6.2 Reelin accumulation in the hippocampus and the ProS-CA1 region in sAD**

We previously reported that peri-plaque Reelin aggregates are evident in the CA2-3 region and the molecular layer of the dentate gyrus (MLDG) in a subset of sAD cases. A representative example is provided in Panel A. Reelin aggregates were less common and less prominent in the ProS-CA1 region. A representative example showing subtle ProS-CA1 Reelin expression in the same sAD case is shown in Panel B.

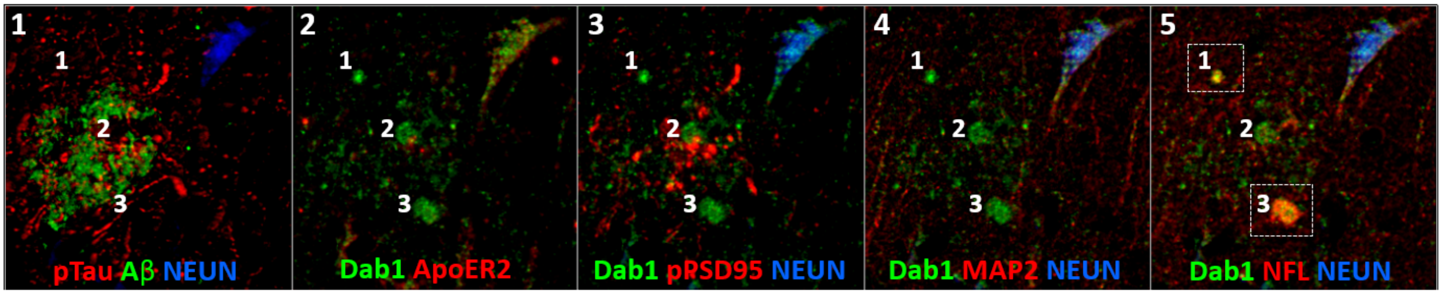


**Ext Fig 9.1 Dab1 accumulation within plaque-associated dystrophic axons in temporal neocortex in sAD**
A coronal section of the temporal neocortex from a sAD case was probed with antibodies targeting Dab1, MAP2, NFL, NEUN, pTau, pPSD95, and Aβ (see **Suppl Table 3**). Dab1 accumulated within three prominent globular structures in the vicinity of an Aβ-labeled neuritic plaque. Two of these three globular Dab1-expressing structures colocalized with NFL-labeled dystrophic axons (demarcated by rectangles in panel 5). None of the three Dab1-expressing structures colocalized with MAP2, pTau or pPSD95.

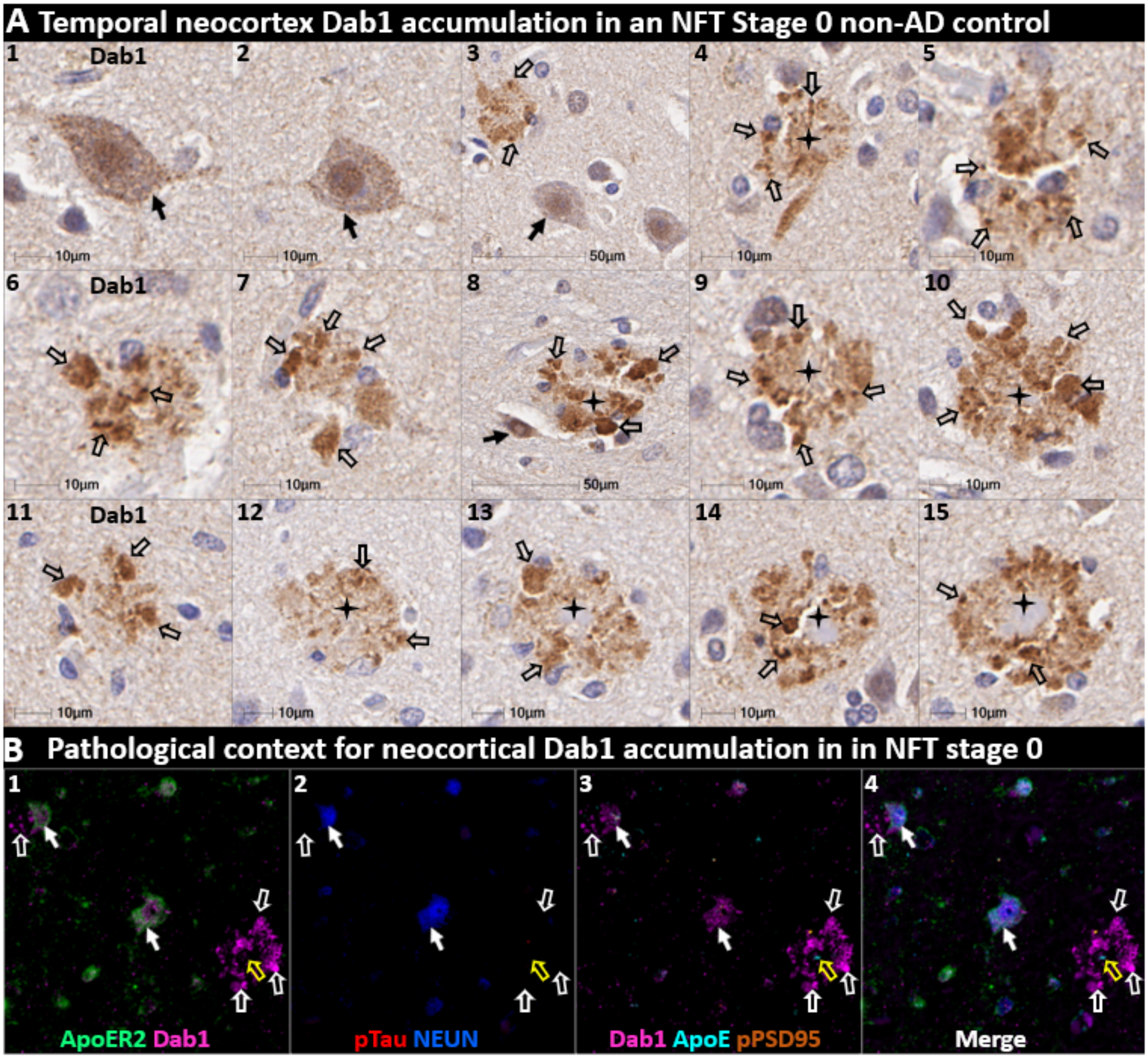


**Ext Fig 9.2 Pathological & cytoarchitectural context for neocortical Dab1 accumulation in NFT stage 0**
Single-target IHC (**$A_{1\text{-}15}$**) of temporal neocortex in a non-AD control with Aβ plaques but no overt pTau pathology (Thal phase 3, NFT stage 0) revealed globular Dab1 accumulations that were most prominent in L5 and L3. Dab1 accumulated in the vicinity of plaque-like structures (designated with stars in **$A_{4,\ 8\text{-}10,\ 12\text{-}15}$**). Dab1 expression was evident in a subset of pyramidal neurons (solid arrows in **$A_{1\text{-}3,\ 8}$**). pTau and $pPSD95_{Thr19}$ were minimally expressed in serial sections. MP-IHC (**$B_{1\text{-}4}$**) revealed that Dab1 accumulated in ApoER2-expressing neurons (white arrows) and that some globular Dab1 accumulations (open white arrows) were clustered around an ApoE-enriched central core (open yellow arrows).

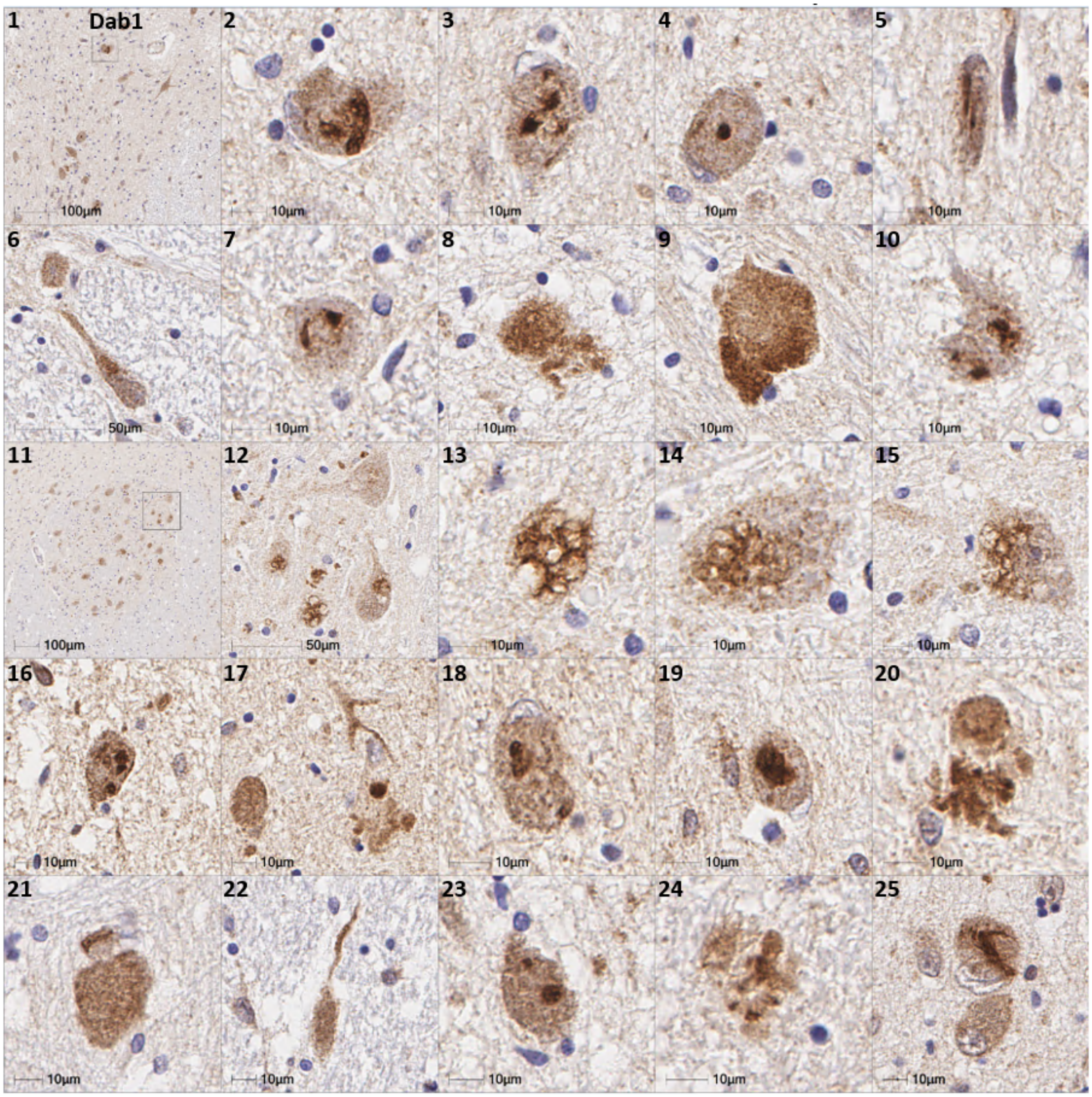


**Ext Fig 10.1. Intra-neuronal Dab1 inclusions in the locus coeruleus and raphe nucleus in sAD**
This figure depicts a variety of morphologies observed for intraneuronal accumulation of Dab1 in pontine LC-PC complex and raphe nucleus in four sAD cases.

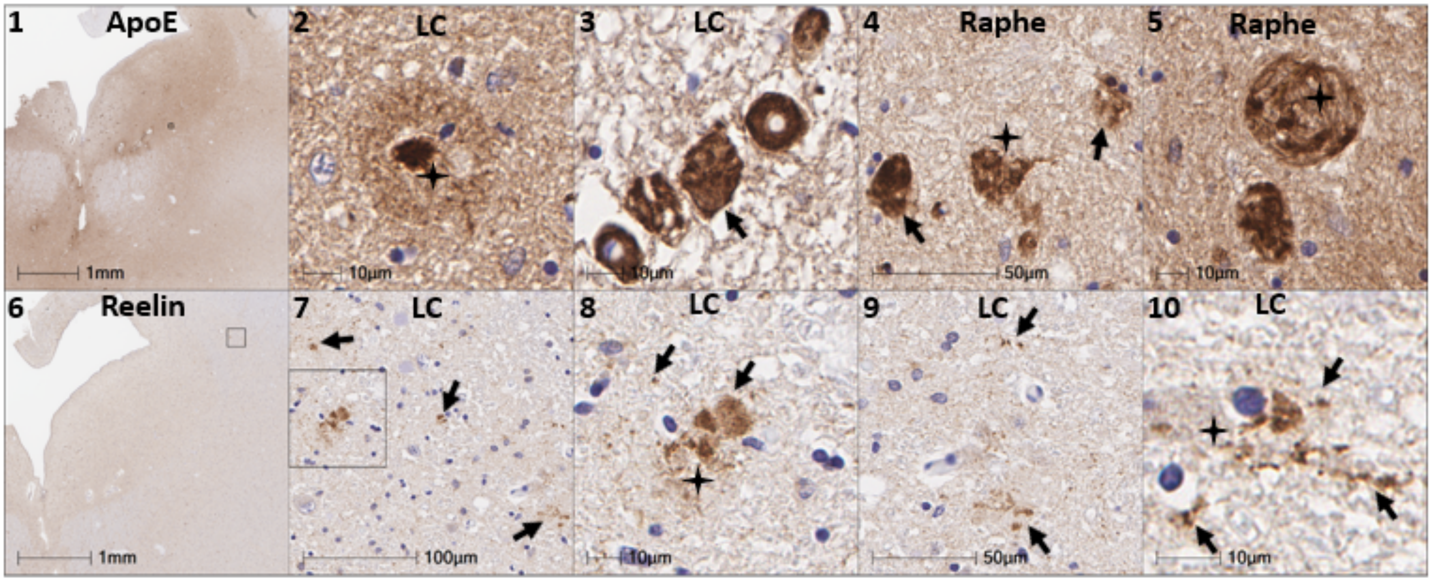


**Ext Fig 10.2. Extracellular accumulations of ApoER2 ligands in locus coeruleus & raphe nucleus in sAD**
Extracellular accumulations of ApoE were observed in many sAD cases (Panels 1-5). ApoE accumulated in both plaque-like structures (Panel 2) and the walls of blood vessels (Panel 3). Extracellular accumulations of Reelin were observed in a subset of sAD cases (Panels 6-10) but were not evident in most cases.
